# Design and approval of the nutritional warnings’ policy in Peru: Milestones, key stakeholders, and policy drivers for its approval

**DOI:** 10.1101/2022.09.12.22279683

**Authors:** Diez-Canseco Francisco, Victoria Cavero, Juan Álvarez-Cano, Lorena Saavedra-Garcia, Lindsey Smith Taillie, Francesca Dillman Carpentier, J. Jaime Miranda

## Abstract

**Background:** Nutritional warnings are used as a public health strategy to prevent increases in obesity prevalence. Peru approved in 2013 and implemented in 2019 a Law requiring nutritional warnings on the marketing and packaging of processed foods high in sugar, sodium, saturated fat, and containing transfat. The complexity behind the implementation of this set of policies over six years provide unique learnings, essential to inform the obesity prevention context, especially when facing strong opposition from powerful stakeholders such as the food industry.

**Aims:** Describe milestones and key stakeholders’ roles and stances during the nutritional warnings policy design in Peru; and identify and analyze the main drivers of policy change that explain its approval.

**Methodology:** In 2021, interviews were conducted with 25 key informants, advocates and opponents of the policy, closely involved in its design. Interviews were analyzed using the Kaleidoscope Model as a theoretical framework. Relevant policy documents and news were also analyzed.

**Results:** Milestones for this policy were the approval of the Law, Regulation, and Manual. Policy supporters were mainly from the Congress, civil society organizations, and Health Ministers; whereas opponents came from other parties in the Congress, ministries linked to the economic sector, the food industry, and media. Across the years, warning’s evolved from a single text, to traffic lights, to the approved black octagons. Main challenges included the strong opposition of powerful stakeholders; the lack of agreement for defining the appropriate evidence for nutritional warning parameters and design; and the political instability of the country. Based on the Kaleidoscope Model, the policy successfully targeted a relevant problem (unhealthy eating decisions) and had powerful advocates who effectively used focusing events to reposition the warnings in the policy agenda across the years. Negotiations weakened the policy but led to its approval. Importantly, government veto players were mostly in favor of the policy, which enabled its final approval despite the strong opposition.

**Conclusions:** Despite the strong opposition faced and technical and political difficulties to define the best parameters and warnings’ design, Peru’s nutritional warnings policy was approved. Lessons learned are essential to inform similar and related prevention policies in Peru and elsewhere.

## I. INTRODUCTION

The prevalence of overweight and obesity in many Latin American countries is increasing more than one percentage point per year (1). This rise has been associated with a greater availability of ultra-processed foods with high contents of sodium, sugars, and saturated fats, which increase the risk of obesity and related non-communicable diseases (NCDs) (2). One strategy to address this problem is the use of nutritional warnings in processed foods’ front-of-package (FoP) labels, which have been recently implemented in Latin American countries, such as Chile, Peru, Uruguay, and Mexico (3). These nutritional warnings inform the public about the high content of nutrients of concern, such as sugar, sodium, and fats (3).

Peru has a long history of undernutrition, but in the last decades, childhood overweight and obesity increased from 25% in 2007 to 38% in 2018 (4). To address this problem, in 2013 Peru launched the Healthy Eating Law (Law 30021), which included the use of nutritional warnings on the processed foods FoP labels and their publicity (5). However, the warnings had to overcome multiple barriers and did not enter into force until six years later, a larger process compared to other countries in the region, such as Chile (6) or Uruguay (7). Indeed, in contrast to Chile (8) or Mexico (9, 10), whose policies processes have been studied and their main challenges, successes, and potential improvements described, the policy in Peru has received little attention. Importantly, Peru included the warnings into a regulation that makes it mandatory for the food industry to comply, and which is joined by other initiatives to promote healthy eating, making it relevant for other low- and middle-income countries (LMICs) who might implement this type of policy.

We performed a qualitative study to analyze the nutritional warnings’ policy design process, using the Kaleidoscope Model of change in agricultural and nutrition policies (11). This model was designed to be used in LMICs, such as Peru, and provides a framework for analyzing why policy-related changes occur; for example, the implementation of a new policy. The model is based on five stages of the political process: agenda setting, design, adoption, implementation, and evaluation and reform, and propose a series of hypotheses to explain the success of a given policy.

In this paper, we describe the milestones and key stakeholders’ roles and stances during the design and approval of the nutritional warnings’ policy in Peru (2013–2019) and analyze the policy drivers that might explain why this policy was finally designed and approved, despite the strong opposition that it faced over the years. To do so, we used the first three stages of the Kaleidoscope Model (agenda setting, design, and adoption), which entails a total of nine determinants and hypotheses, as shown in Table 1:

**Table 1.**
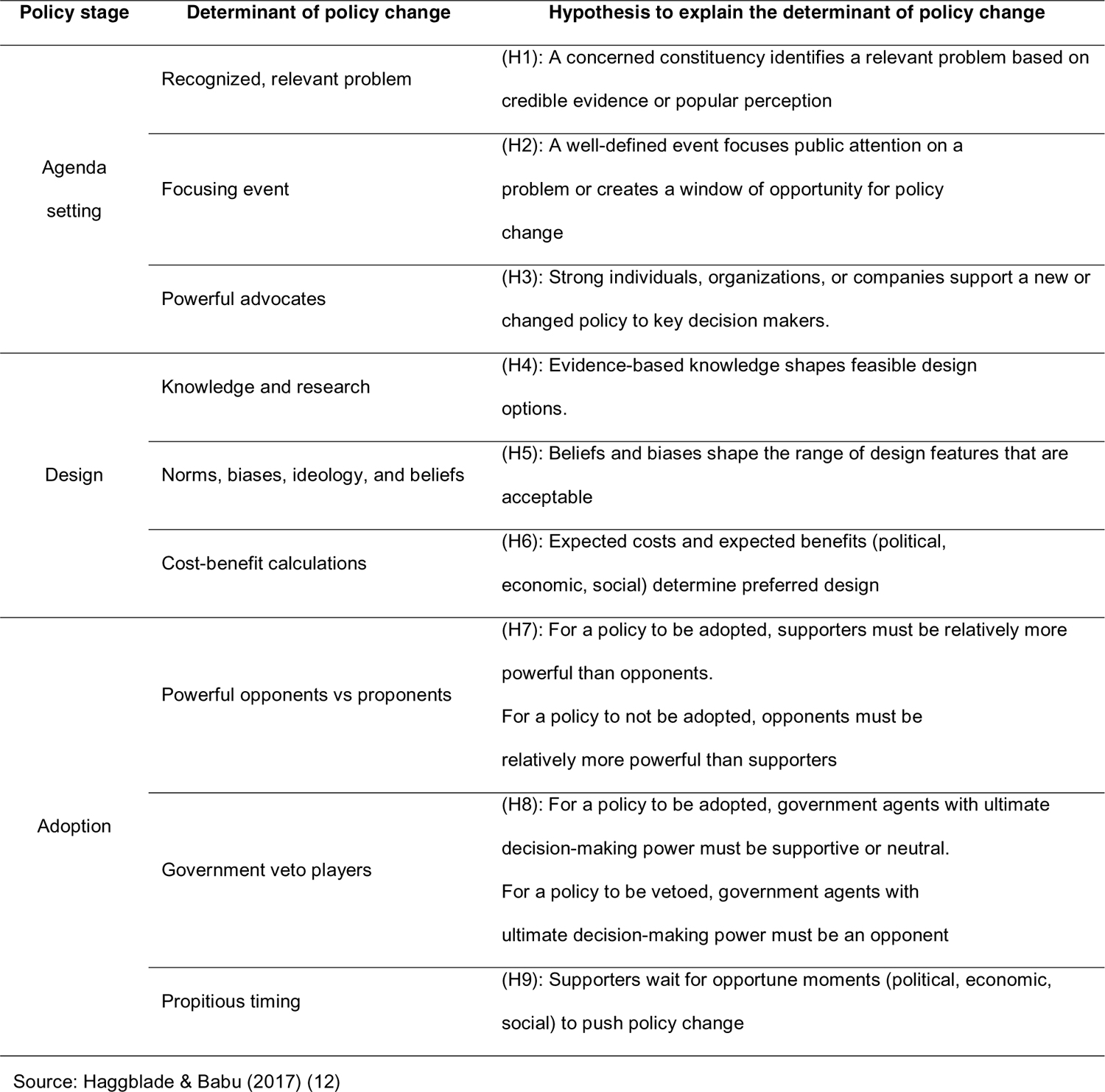
Components of the Kaleidoscope Model

## II. Methods

### 1. Context about the nutritional warnings policy in Peru

The nutritional warnings policy in Peru comprises three main documents: The Law 30021, its Regulation, and the Nutritional Warnings Manual. In May 2013, the Peruvian Congress enacted Law 30021 (5) to reduce children and adolescents’ overweight and obesity and prevent NCDs. The Law contains six strategies, including the use of nutritional warnings (later designed as black octagons) on the processed foods FoP labels and their publicity. To implement the strategies, the Law delegated the Ministry of Health (MoH) to design a regulation, based on recommendations of the World Health Organization (WHO) or the Pan American Health Organization (PAHO), to establish parameters for maximum levels of sugar, sodium, saturated fat, and trans-fat for processed foods and beverages. If one or more of these parameters were surpassed, the corresponding nutritional warning would be used. The MoH had 60 days to prepare the regulation, which entailed a policy document to specify the Law’s contents. The Law, however, was not approved until four years later, in June 2017. This Regulation (13) specified two implementation dates of the warnings, with stricter nutritional parameters for the second one; and commissioned the MoH the preparation of a “Nutritional Warnings Manual” to detail the warnings’ design (e.g., shape, size, color, etc.). The Manual (14) was approved in June 2018, and the implementation of the nutritional warnings begun in June 2019, followed by the introduction of the stricter parameters in September 2021.

The design and approval of these three documents (Law, Regulation and Manual) faced strong opposition from the food industry and from powerful stakeholders in different sectors and levels of the government, e.g., political parties in the Congress, or Ministries linked to the economic sector. However, the policy also had highly committed advocates in Congress, civil society organizations, PAHO, and decision makers in the MoH and central government.

### 2. Study design

A qualitative study, consisting of semi-structured interviews with stakeholders, was performed during 2021 to collect first-hand information about the process of designing and approving of the nutritional warnings policy in Peru.

### 3. Participants

The informants were stakeholders, from different backgrounds and sectors, closely involved in the design, implementation and/or monitoring of the nutritional warnings policy in Peru. The selection of participants aimed to invite up to 25 key informants, advocates and opponents of the policy, representing: (1) local policymakers (2) politicians, (3) international organization representatives, (4) civil society advocates, (5) industry and media representatives, and (6) researchers. Some participants filled more than one role at different time-points (i.e., policymaker and civil society advocate). A purposive sampling strategy was used to select key informants who could provide detailed descriptions and valuable insights about the policy process from different perspectives. We interviewed 25 informants, as shown in Table 2.

**Table 2.** Total of informants per sector

| Sector | Informants |
| --- | --- |
| <u>Policymakers</u>: Former ministers, responsible of designing the Regulation's and Manual's drafts | 9 |
| <u>Members of Congress</u>: Supporting and opposing the policy (based on votes, bills to modify the Law) | 3 |
| <u>International organization representatives</u>: Former and current representatives | 3 |
| <u>Civil society advocates</u>: Former and current representatives involved in the policy | 4 |
| <u>Industry and media representatives</u>: Representatives from associations and from individual companies | 3 |
| <u>Researchers</u>: Market studies' researchers and master students | 3 |
| **Total** | **25** |

### 4. Data collection tools

One semi-structured interview guideline was developed and organized into seven sections comprising questions about the design, implementation, and audit of the nutritional warnings policy in Peru. This comprehensive guideline was adapted to each interviewee, prioritizing those topics closer to their experience during the policy design, implementation, and audit. New queries emerged during the data collection and were included in the following interviews if relevant.

Additionally, a rapid search using Google engines was conducted to retrieve news and policy documentation about the nutritional warnings policy to contextualize the interviewees’ information and better understand the main policy contents and events. Search terms included “nutritional warnings”, “octagons”, “healthy eating law”, and “Law 30021”. News were selected from Jan 1^st^, 2011 to June 30^th^, 2018, when the Manual was approved. Likewise, and using the Peruvian Government official website (www.gob.pe), the main policy documentation (i.e., the approved Law, Regulation and Manual, as well as previous and later drafts) were collected.

### 5. Procedures

Interviews were conducted in Spanish between January and September 2021 through video calls and were facilitated by four researchers involved in the project since its conception. All interviews were audio recorded after obtaining informants’ consent, and their average duration was 84 minutes (range: 53–118 minutes). When necessary, more than one session was arranged to cover all the main topics of the interview guideline. News and policy documentation were read and summarized by one researcher involved in the interview process.

### 6. Data analysis

Interview recordings were transcribed and a directed qualitative content analysis was performed (15) using ATLAS.ti 9. All interviews were double coded by two trained researchers, who lead or attended the interviews.

An initial codebook was developed based on the interview guideline’s topics, with the aim of capturing essential milestones of the policy design and approval (i.e. approval of policy documents about the nutritional warnings), the role and stances of main stakeholders, and variables from the Kaleidoscope Model (e.g., focusing event,). Before coding, the team met to standardize the procedure and understanding of the codes to be used. Along the coding process, emerging codes were discussed to decide if they would be added to the codebook. Disagreements were solved by discussions with another member of the research team. The final codebook had 124 codes (Supplementary Material). This manuscript mainly presents the analysis of codes referring to the policy agenda setting, design, and adoption, from 2013 to 2019. The implementation and audit processes are described elsewhere. Additionally, the most relevant policy documentation and news about the nutritional warnings’ design and approval were used to contextualize and triangulate the informants’ discourses and to prepare a timeline of main events.

### 7. Ethical considerations

The study was approved by Institutional Review Boards at Universidad Peruana Cayetano Heredia and University of North Carolina at Chapel Hill. All interviewees gave informed consent. Some signed a virtual form while others gave oral consent during a recorded phone call.

## III. Results

Results are divided in two sections. First, we present the milestones during the nutritional warnings’ design by describing the processes of designing and approving the Law 30021, its Regulation, and the Manual (Table 3), as well as the role of key stakeholders along the years (Figures 1-3). We also outline how the nutritional warnings’ parameters and design were defined along the years and summarize the main challenges faced during these processes. Based on this narrative, in our second section we use the Kaleidoscope Model to analyze how and why this policy was ultimately approved, despite the long periods and strong opposition. Hypotheses derived from the key contextual factors, actions, and actors associated with the model’s stages of agenda-setting, design, and adoption (see Table 1) are addressed in this second section.

**Figure 1.**
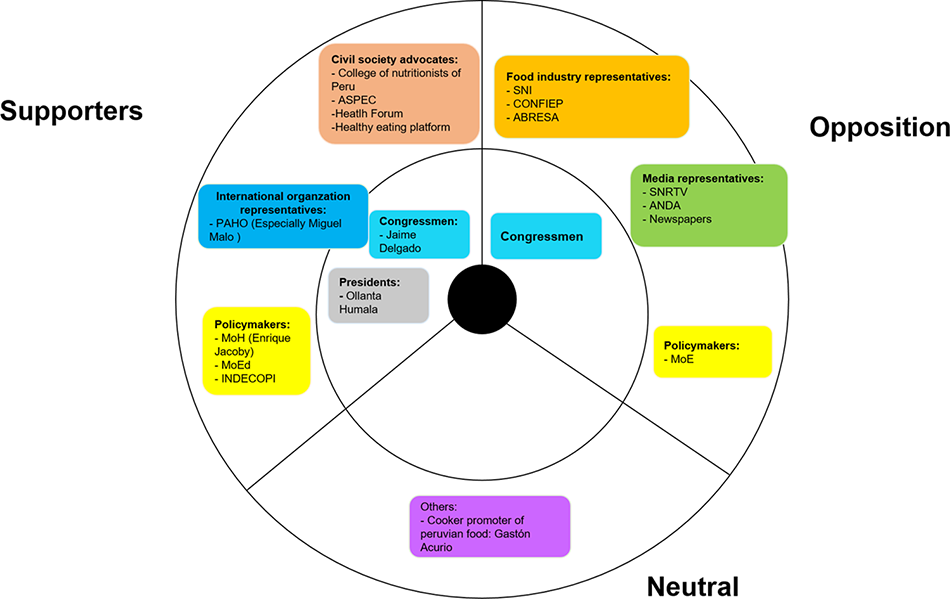
Stances of key stakeholders during the Law 30021’s design and approval (2012–2013) Stakeholders in the inner circle are those with higher power in the decision-making process <u>Abbreviations</u>: Ministry of Health (MoH), Ministry de Economy and Finances (MEF), Ministry de Education (MoE), Ministry of Justice (MoJ), Ministry of Social Inclusion (MoS), Ministry of Foreign Trade and Tourism (MoF), Ministry of Agriculture (MoA), Ministry of Production (MoP), National Radio and Television Society (SNRTV), National Advertisers Association (ANDA), National Confederation of Private Entrepreneurial Institutions (CONFIEP), National Society of Industries (SNI), Non-alcoholic Beverage and Soft Drink Industry Association (ABRESA), Peruvian Association of Consumers and Users (ASPEC)

**Figure 2.**
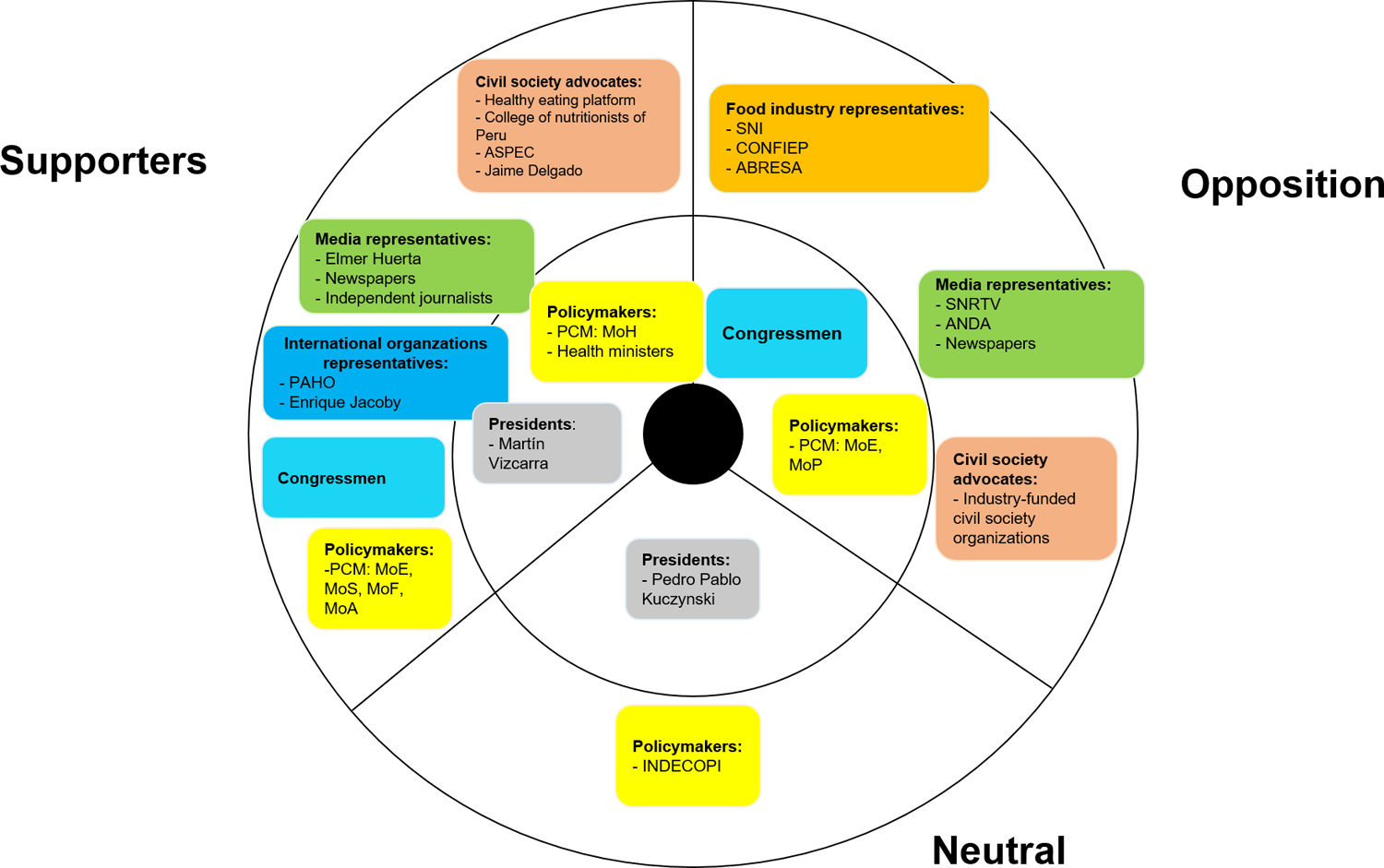
Stances of key stakeholders during the Regulation’s design and approval (2014–2017) Stakeholders in the inner circle are those with higher power in the decision-making process <u>Abbreviations</u>: Ministry of Health (MoH), Ministry de Economy and Finances (MEF), Ministry de Education (MoE), Ministry of Justice (MoJ), Ministry of Social Inclusion (MoS), Ministry of Foreign Trade and Tourism (MoF), Ministry of Agriculture (MoA), Ministry of Production (MoP), National Radio and Television Society (SNRTV), National Advertisers Association (ANDA), National Confederation of Private Entrepreneurial Institutions (CONFIEP), National Society of Industries (SNI), Non-alcoholic Beverage and Soft Drink Industry Association (ABRESA), Peruvian Association of Consumers and Users (ASPEC)

**Figure 3.**
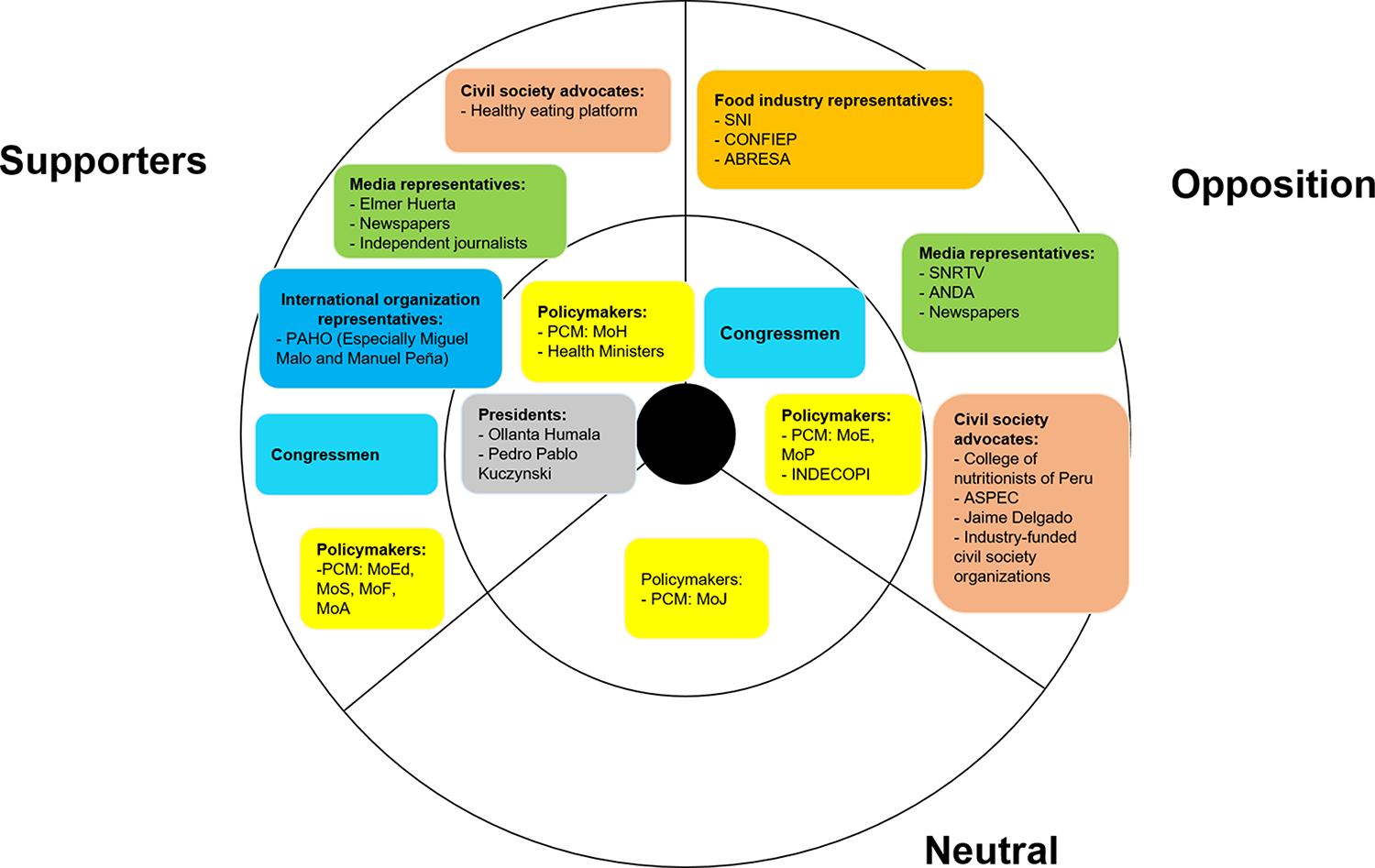
Stances of key stakeholders during the Nutritional Warnings Manual design and approval (2017–2018) Stakeholders in the inner circle are those with higher power in the decision-making process, <u>Abbreviations</u>: Ministry of Health (MoH), Ministry de Economy and Finances (MEF), Ministry de Education (MoE), Ministry of Justice (MoJ), Ministry of Social Inclusion (MoS), Ministry of Foreign Trade and Tourism (MoF), Ministry of Agriculture (MoA), Ministry of Production (MoP), National Radio and Television Society (SNRTV), National Advertisers Association (ANDA), National Confederation of Private Entrepreneurial Institutions (CONFIEP), National Society of Industries (SNI), Non-alcoholic Beverage and Soft Drink Industry Association (ABRESA), Peruvian Association of Consumers and Users (ASPEC)

**Table 3.**
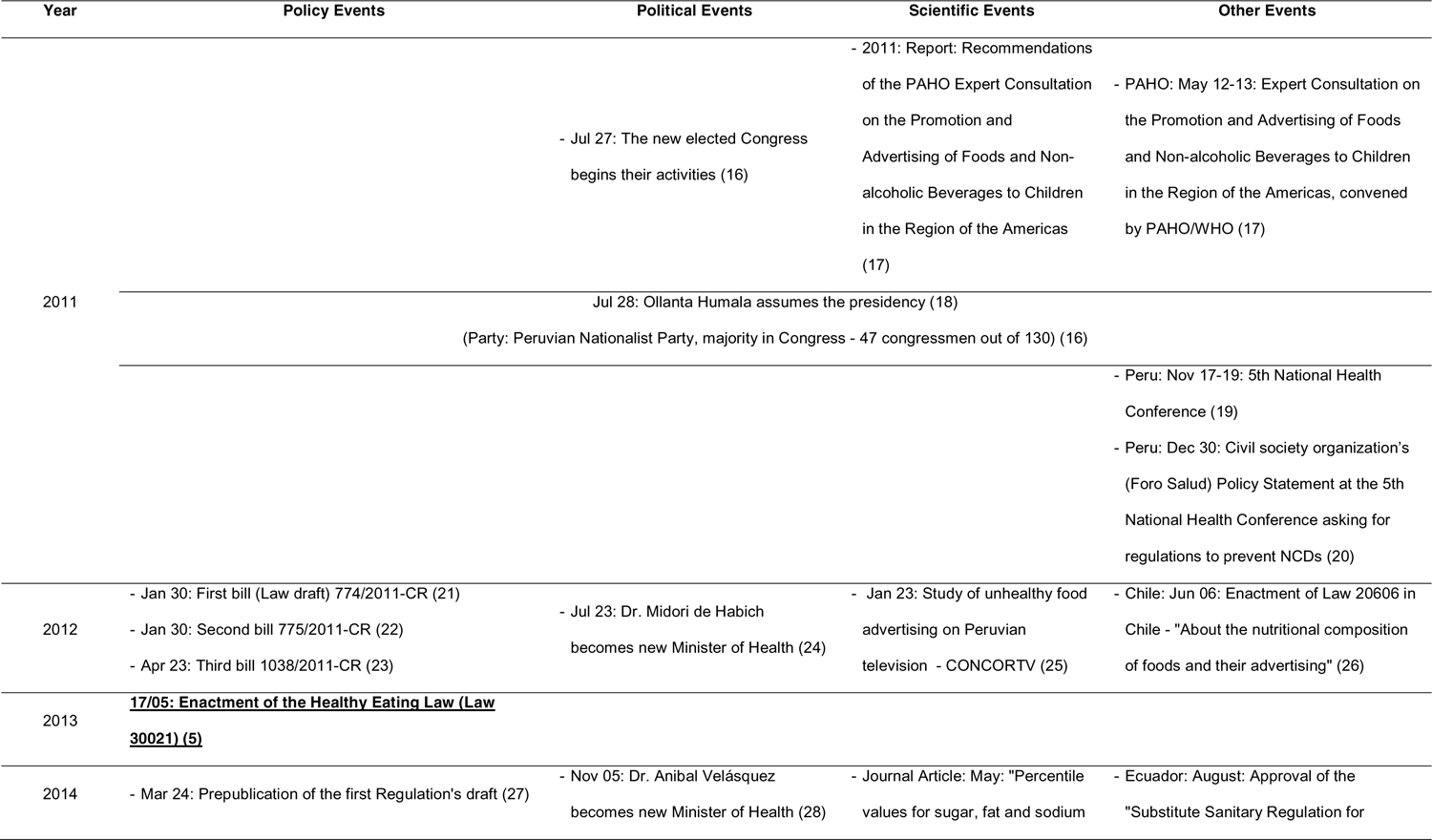
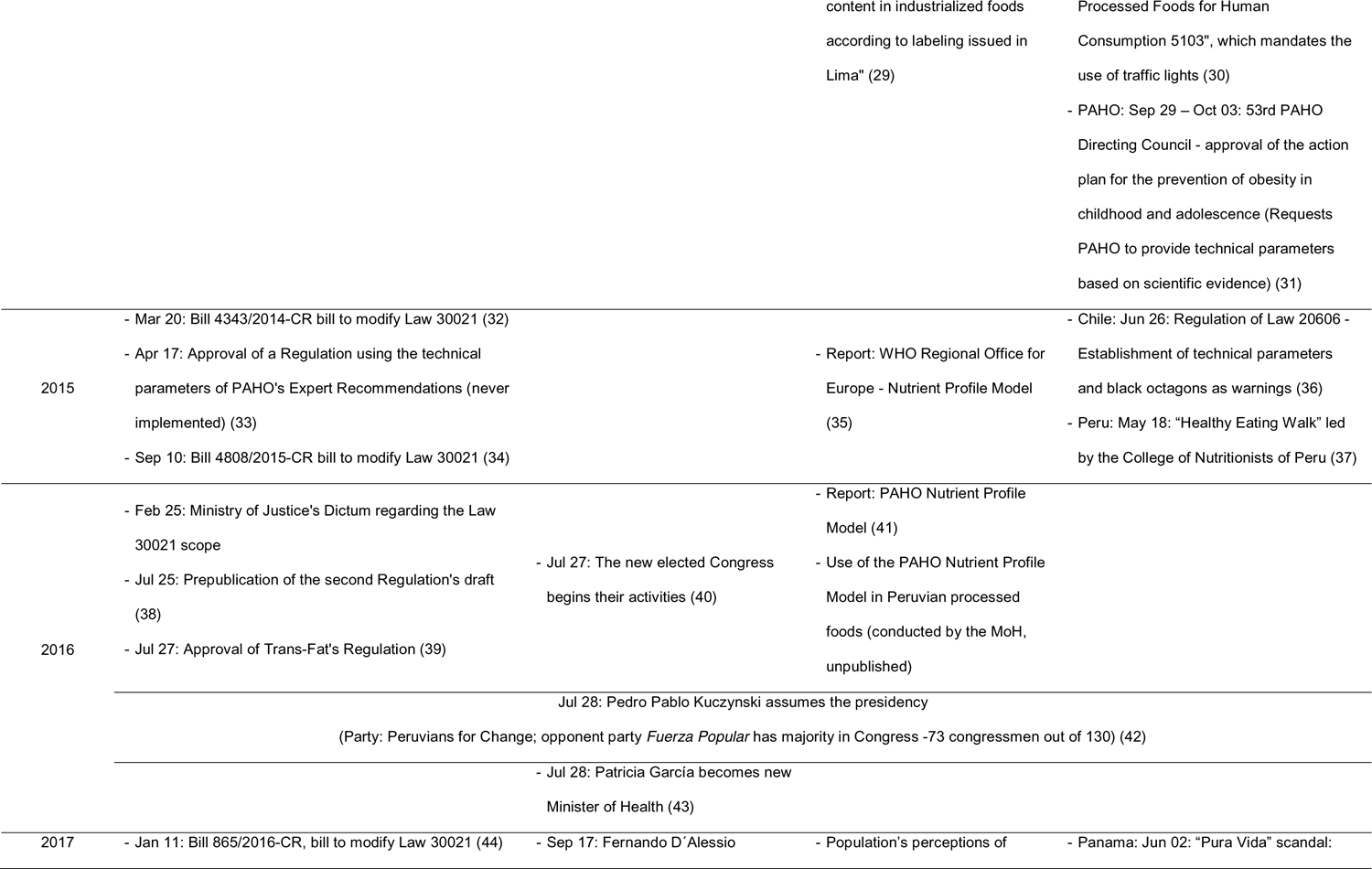
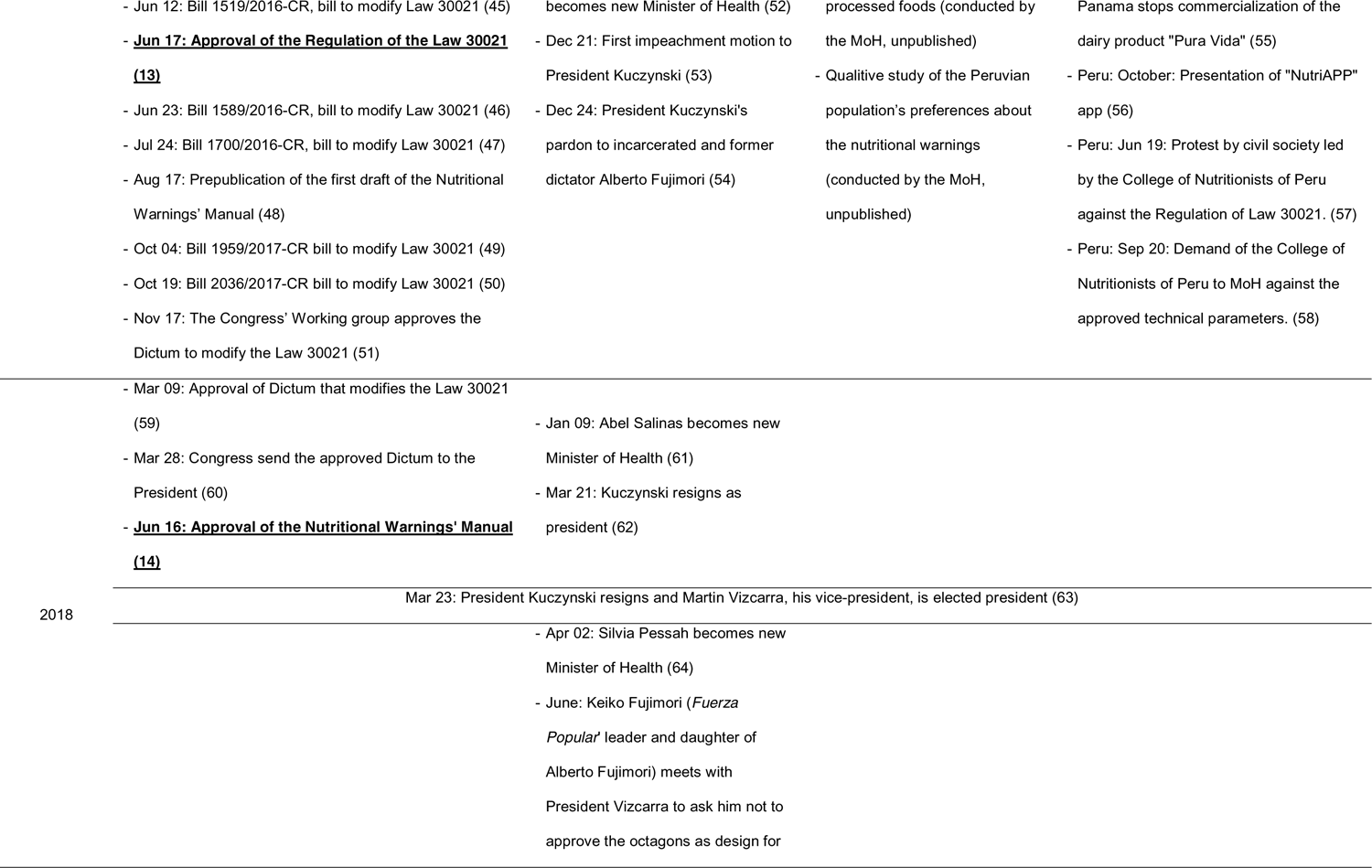
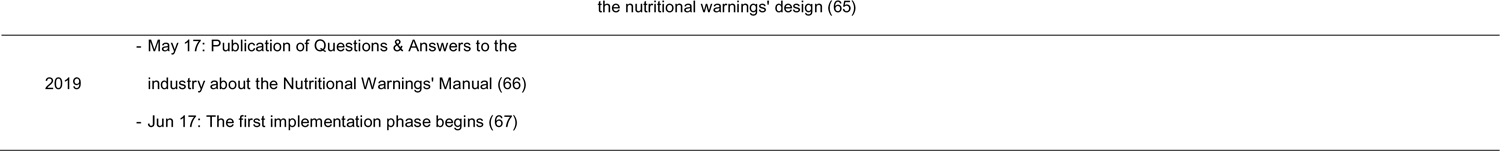
Timeline of main events related to the Nutritional Warnings Policy’s design and approval in Peru, 2011-2019.

## 8. Milestones during the nutritional warnings’ design

### 8.1. In 2013, nutritional warnings in FOP labels and publicity are included in the Law 30021

#### 8.1.1. Summary of how the Law was designed and approved

According to our informants, the nutritional warnings in Peru aimed to reduce obesity and promote healthy eating habits in children and adolescents by 1) informing the population about the high content of nutrients of concern in processed food; 2) encouraging the consumption of healthier options (with less or no warnings); and 3) encouraging the food industry to improve the nutritional content of their products. These warnings reached de Peruvian policy agenda as one of the six components of the “Healthy Eating Law” (Law 30021) (5).

This Law was the result of three bills (Law drafts) designed by Peruvian congressmen from different parties between January and April 2012 (21–23). These bills were merged into one single text and discussed in the Congress for several months, when the text was constantly edited mainly based on critics from media and food industry representatives but also on some recommendations from public institutions, civil society organizations, and PAHO (68). Despite the strong opposition and influence of food industry and media representatives, congressmen in favor of the Law were majority than those against, which enabled its enactment and further endorsement of the country President Humala (2011–2016), in May 2013. The constant editions removed important elements that many interviewees regretted, but they also commented that it was the best that could be achieved after all the opposition received.

> “The congressman who was in charge told me ‘Ms. that’s what was possible, this is the Law that we were able to do’. (…) So, yes, that left me thinking a lot. I then began to say: “The Law is not perfect, but it is the only thing, the best we have been able to achieve and with this imperfect Law we can now move forward”. (Civil society organization’s representative, SOC-01)

#### 8.1.2. Definition of the nutritional warnings’ parameters in the period of designing the Law

Nutritional warnings required parameters to define the maximum nutrients’ content that a processed product could have to avoid the warning in its FOP label and publicity. The third bill (before the Law) (23) proposed using a set of parameters produced during an PAHO expert consultation in 2011 (17), which established maximum contents of sugar, fat, sodium, and trans-fat for solid (grams) and liquid (milliliters) processed foods (Table 4). However, opponents argued that these parameters were based on experts’ opinions and not on an official PAHO document nor scientific evidence. This argument was supported by the National Institute for the Defense of Free Competition and the Protection of Intellectual Property (Indecopi) (68), the governmental institution in charge of auditing the nutritional warnings’ implementation. Thereby, the Law did not include any parameter but instead commissioned the MoH to define them in a further Regulation (5). This Regulation should be published in no more than 60 days and parameters should be “based on recommendations from WHO/PAHO”.

**Table 4.**
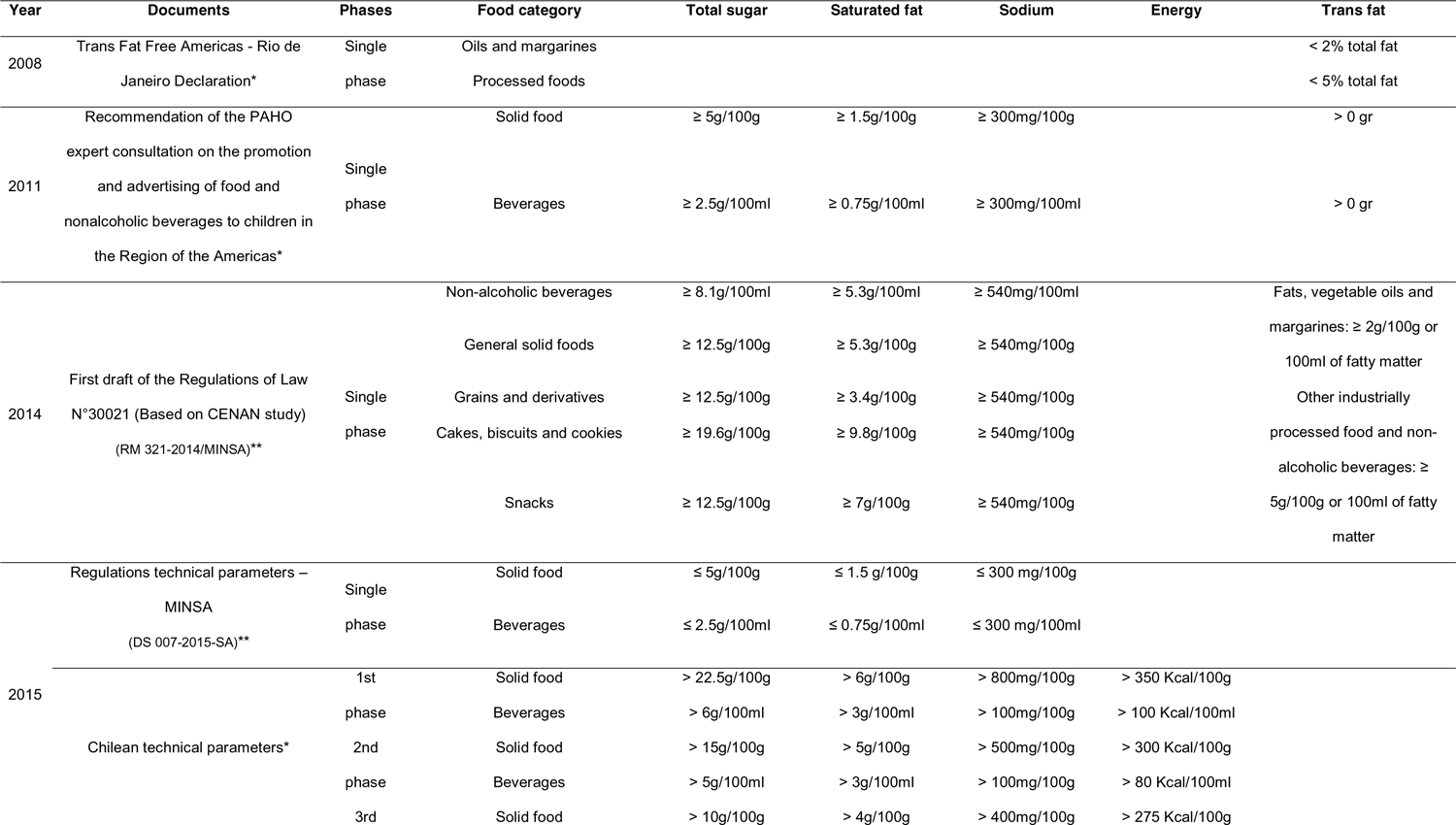
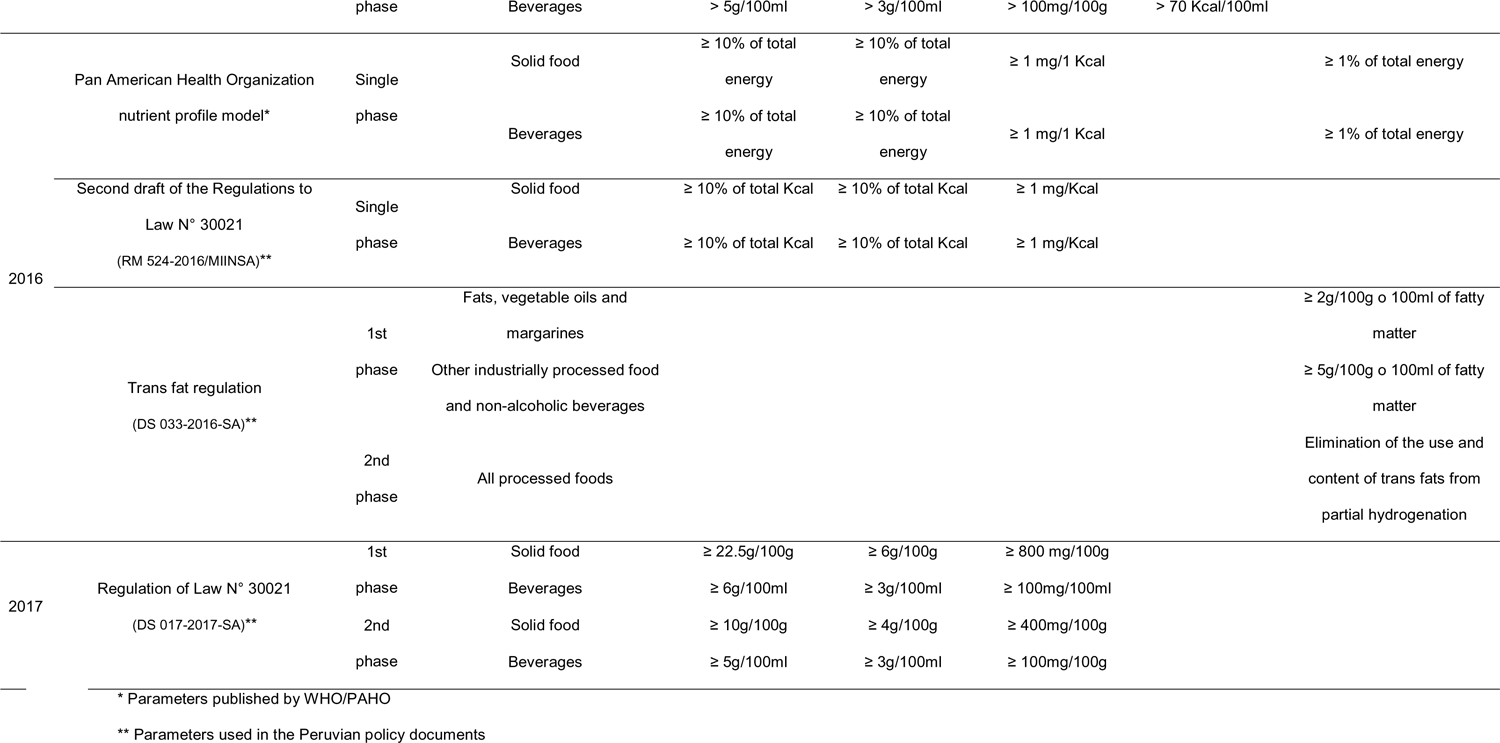
Timeline of parameters for maximum levels of sugar, saturated fat, sodium, and trans-fat in processed foods and beverages, including those proposed by WHO/PAHO and those used in the nutritional warnings’ policy documents

#### 8.1.3. Proposed nutritional warnings in the period of designing the Law

The Law did not mention any specific design for the nutritional warnings but only the following texts: “High in [sodium/sugar/fat]: Avoid its excessive consumption” and “Contains trans-fat: Avoid its consumption”, specifying that they should be clearly seen in the products’ FoP labels and their publicity (5), similar to the warnings used in tobacco. According to our informants, neither the policy advocates nor opponents proposed any specific design in terms of shape or colors when designing the Law. See Figure 4 to appreciate the different designs proposed across time and Table 5 to understand how the warnings design changed from traffic light label to a warning label.

**Figure 4.**
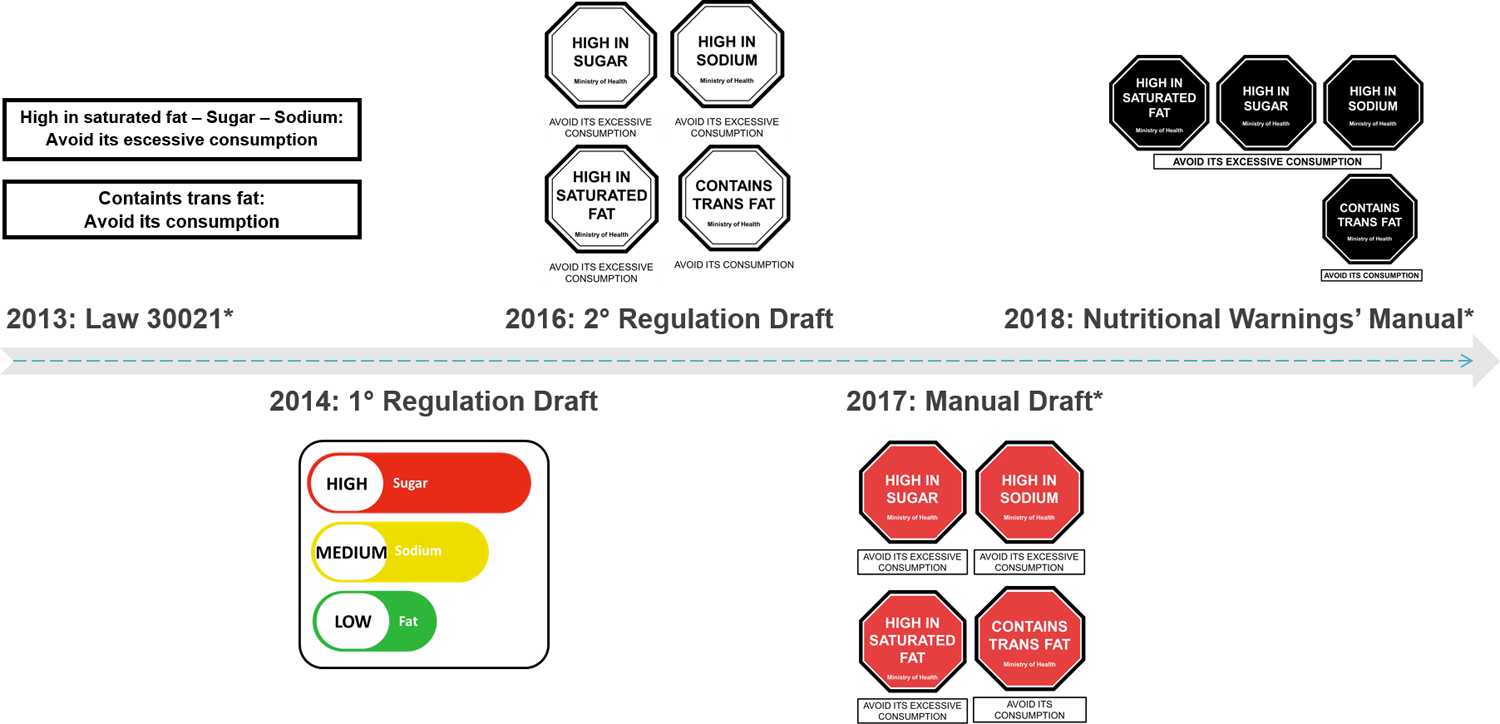
Nutritional warnings’ designs proposed during the main policy documents design *Only these designs were published in official documents, The Law specified the text for the nutritional warnings, similar to the ones used in tobacco. When designing the first Regulation Draft, its authors suggested using the traffic lights design, similar to the ones being used in Ecuador at that time. Since the second Regulation Draft, the authors suggested using the octagons design, but with some differences, such as color and shape.

**Table 5.**
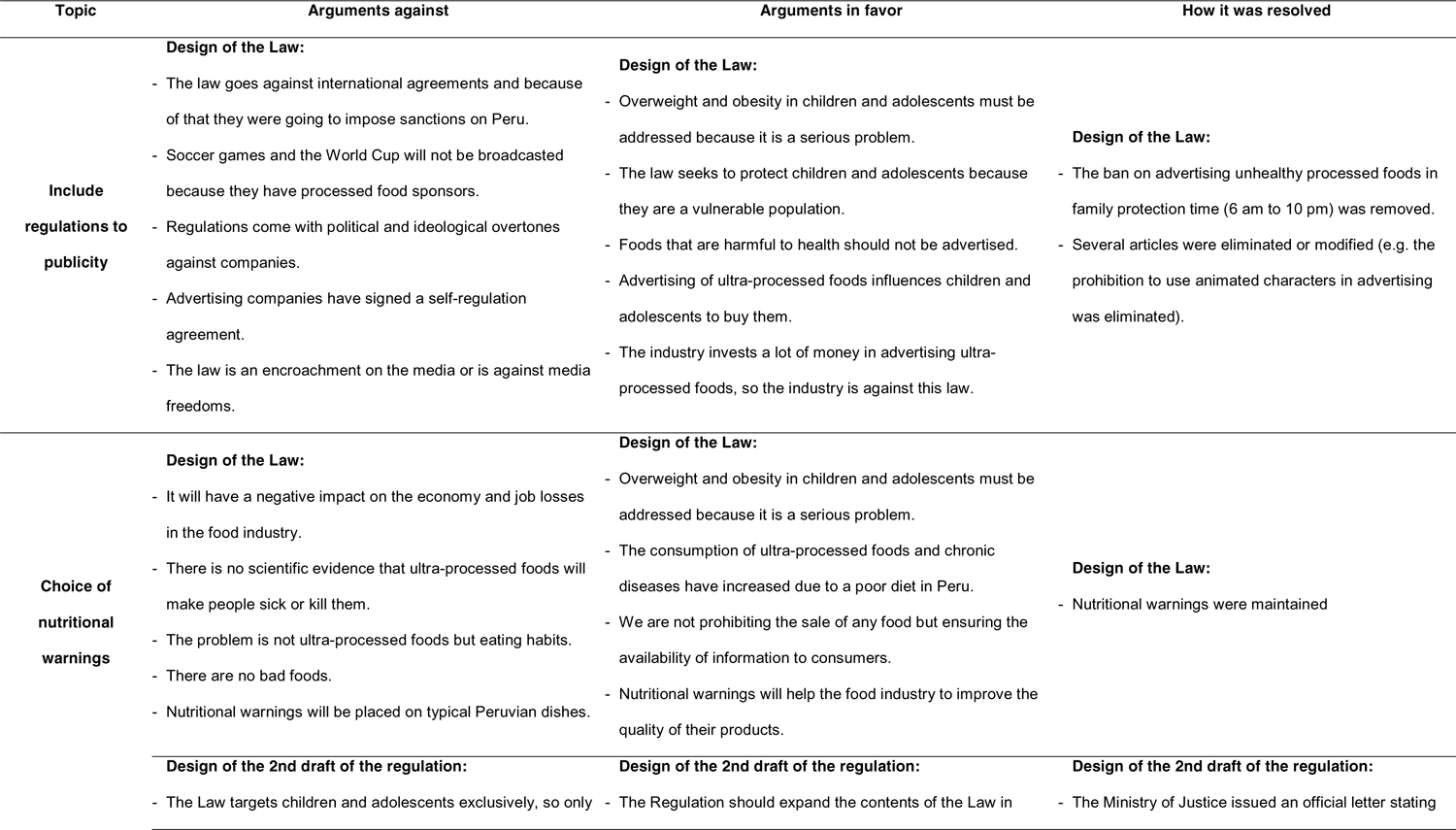
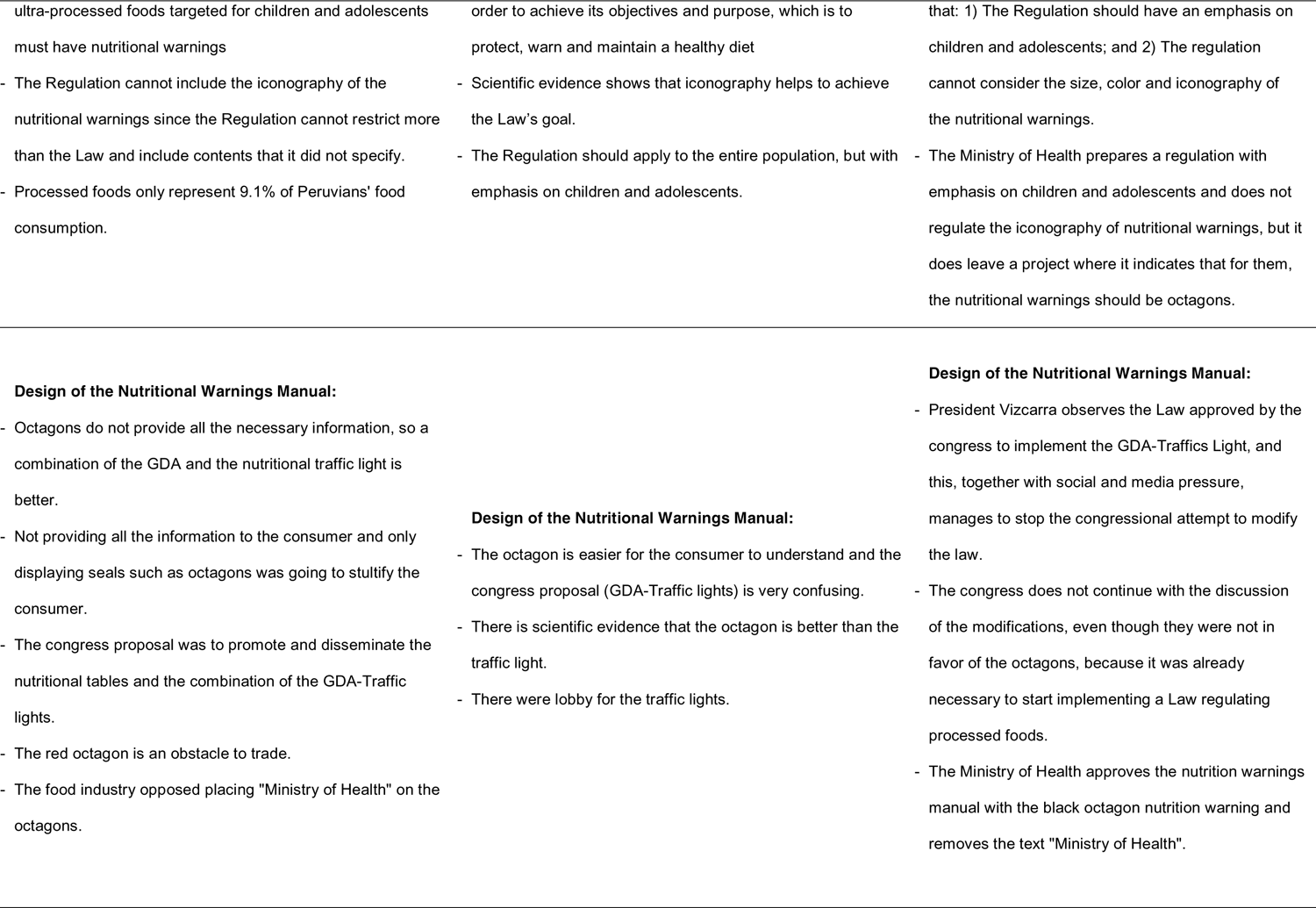
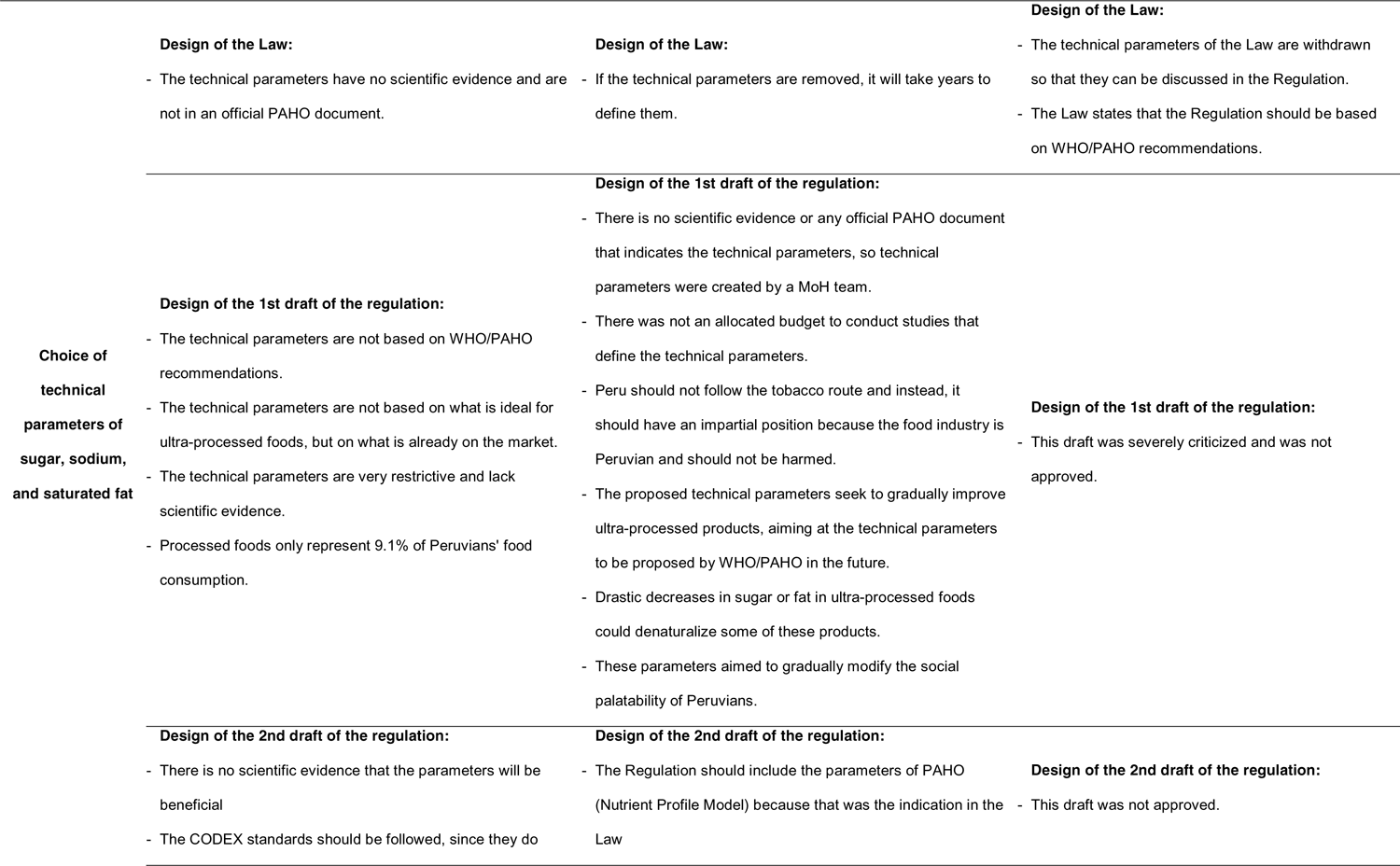
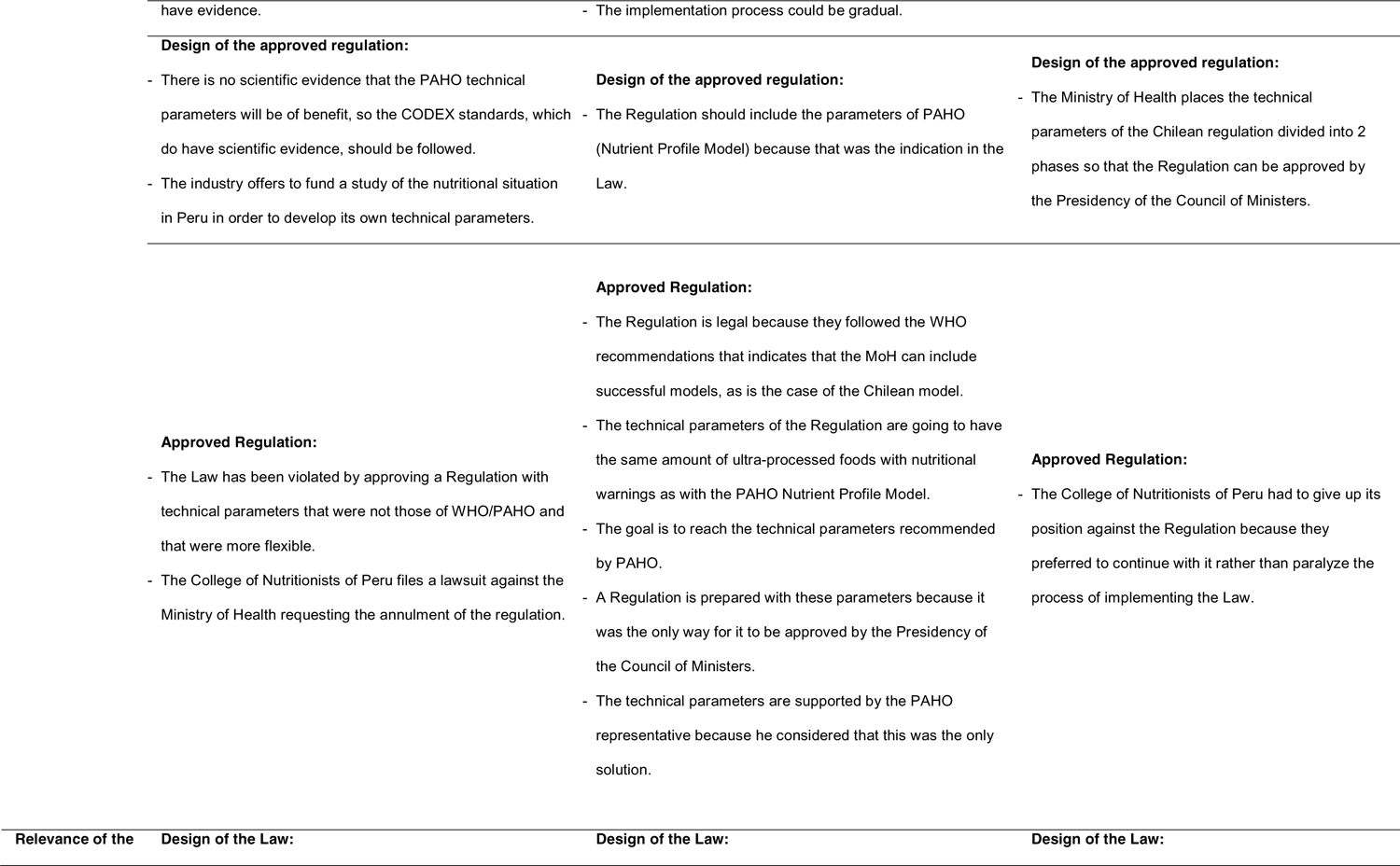
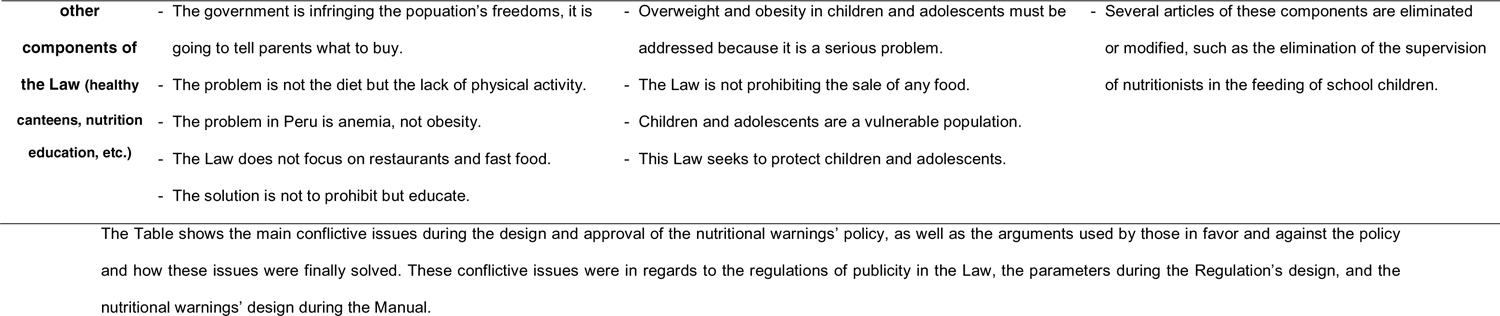
Main conflictive issues among supporters and opponents of the nutritional warnings

#### 8.1.4. Main challenges faced when trying to approve the Law 30021

Main challenges in this period were 1) the strong opposition of food industry and media representatives to some aspects of the Law and 2) the little scientific evidence to define the nutritional warnings’ parameters. Arguments used by those in favor and against the warnings can be found in Table 5. Importantly, some informants said that during this period, opponents were mainly focused on the regulations to publicity (another component of the Law) rather than in the nutritional warnings, which might have eased including the warnings in the Law. However, this strong opposition risked the approval of the Law.

> “People from the Society of Radio and TV said: ‘You know what, this project can’t be approved with that [regulations to publicity]. We are not interested if you put labels, whatever… [but] don’t mess with publicity’. The same day, someone from the Fujimori’s party, comes and says, ‘If you don’t take that out, that article, don’t even count with our votes’. And then others from another party, who were supposedly my allies, ‘If you don’t take that out, there is no, there is no Law’. And I’m like ‘Wow, how much power!’” (Congress member 2011-2016, POL-02).

### 8.2. In 2017, the parameters for the nutritional warnings are finally defined in the Regulation of the Law 30021

#### 8.2.1. Summary of how the Regulation was designed and approved

Contrary to the expectation of having the Law’s Regulation in July 2013, it was finally approved four years later, in June 2017 (13). During these years, the MoH pre-published two Regulation drafts (38, 69) with different parameters for sugar, salt, and saturated fat in each, as shown in Table 4. Informants explained that these drafts had to be pre-published for 90 days to receive opinions from private and public institutions before its official approval. According to our interviewees, for each draft, the MoH assigned two working groups to design the Regulation. The sectoral working group, composed only by the MoH, in charge of defining the nutritional warnings’ parameters; and the multisectoral working group, composed by representatives from different ministries and Indecopi, to design the Regulation for all the Law.

In this same period, some congressmen against the Law proposed bills to modify the Law. Two bills were presented in 2015 (32, 34) and two others in 2017 (44, 45). In 2016, Peru had national elections and a new President and Congress were elected. In this new Congress, the main opposing party to the Law had majority of congressmen, which eased the discussion of the proposed bills to modify the Law. Informants from the MoH commented on the pressure and challenging atmosphere of having to design the Regulation while having the Congress attempting to change the Law. Also in these years, civil society organizations led protest marches and were in news programs to demand the implementation of the Law (37). Our informants believed that thanks to these marches, campaigns, and interviews, the Law achieved public awareness and pressed the MoH to accelerate their work.

In June 2017, a large Peruvian company of dairy products was sanctioned by the government of Panama for advertising a product named “Pura Vida” as milk when it was a dairy mix (a beverage made of milk and vegetable products) (70). This case received great media coverage, which proponents of the Law used as an opportunity to advocate for the Regulation’s approval arguing that consumers should be properly informed about what they are eating, which was the purpose of the nutritional warnings. Civil society organizations led protests and made media appearances, pressing the new government to approve the Regulation (57). On June 15^th^, 2017, two weeks after the “Pura Vida” scandal, the Regulation was finally approved.

> “Without “Pura Vida” we would not have a Regulation. I am completely sure of that.” (Decisionmaker at the MoH, DEC-07)

The approved Regulation used the parameters of Chile and commissioned the MoH to design a Manual with the technical characteristics of the nutritional warnings and to approve it by Supreme Decree in no more than 120 days.

#### 8.2.2. Definition of the nutritional warnings’ parameters in the period of designing the Regulation

Once the Law was approved, the MoH oversaw the design of its Regulation. During all these years, the food industry proposed the daily reference values defined by the CODEX Alimentarius (72), which is a set of guidelines and standards approved by WHO. The MoH, however, did not recommend their use for being based on the nutritional requirements of adults and not children or adolescents, and proposed different parameters along the years.

As shown in Table 4, by 2014 the only available source for parameters were the ones proposed in the PAHO’s expert consultation, which were already rejected by those in favor and against the Law for being opinions and not based on scientific evidence. Thereby, the MoH pre-published a Regulation’s draft with parameters for sugar, sodium, and fat, based on an existing database of 400 processed foods and beverages collected in July 2013 (69). This draft, however, was strongly criticized for using parameters too flexible and for not being based on the WHO/PAHO recommendations, as indicated in the Law.

> “Someone who criticized this Regulation a lot was Manuel Peña [former PAHO representative]. He created a strong pressure when he said in the media that this Regulation was not the one that was [proposed] in the Law, that the values [of this draft] were not the ones specified in the Law” (Decision maker at the MoH, DEC-01)

In 2015, and with a new minister of health, the MoH held several meetings with PAHO in Peru and officially asked for their parameters. Our informants commented that PAHO replied by sending the parameters from the PAHO’s expert consultation of 2011, which was used as evidence by the sectoral commission to publish a Regulation’s draft with these parameters (33), in an attempt to cut off discussions at the multisectoral working group that might have led to the adoption of less stringent parameters.

> “We could not issue a new version of the Regulation if it was not done in a multisectoral working group, and that would have taken a long time. So, we decided that the MoH defines the parameters, which were the most critical point (…) (and) The other things, (for example) the label’s design or the timeframe, all that, was part of the Regulation. But the value would not be in dispute. So, what was avoided was the negotiation of parameters. They would be already established.” (Decision maker at the MoH, DEC-01)

Some months later, Chile issued its own parameters for their nutritional warnings (36), which were more flexible than those of the expert consultation but more stringent than those proposed in Peru in 2014 (Table 4). In February 2016, WHO and PAHO published the Nutrient Profile Model (35, 41), which were much stricter than all the previous ones. In face of this new evidence, the MoH replaced its Regulation’s draft of 2015 and used the parameters of the Nutrient Profile Model for the second draft, pre-published on July 25^th^, 2016, (38) days before President Humala finished his ruling period (28^th^ July, 2016).

The parameters for trans-fat were published separately, on July 26^th^, 2016 (39), as they were prepared by a different MoH group who had already defined them in 2012 (71). The trans-fat pre-publication established their elimination in any processed product using partial hydrogenation after 54 months. Interviewees from different sectors agreed that even though the food industry questioned that zero trans-fat would not be technically feasible, they were mainly in favor of the initiative, possibly explaining why these parameters were easy to approve and are still in force.

> “Trans-fats were easier, right? Because in trans-fat there is a consensus that it has to be zero.” (Decision maker at the MoH, DEC-01)

The new government’s MoH, under the presidency of Kuczynski (2016–2018), received several queries, suggestions, and critiques for the second Regulation’s draft. The MoH hired a group of consultants to attend these comments and prepare a new Regulation. The parameters (based on the WHO/PAHO Nutrient Profile Model) were one of the main conflictive issues among the Presidency of the Council of Ministers (PCM), who had to approve the Regulation (Table 5).

> “The minister [of Health] returned several times with the Regulation in hand saying: ‘No, no, this does not pass in the PCM. The minister of Industry, the prime minister, they don’t want, they say no.’” (International organization’s representative, INT-01)

After the “Pura Vida” scandal, and aiming to overcome the PCM’s resistance, the MoH adopted the Chilean parameters and the Regulation was rapidly approved, in June 2017, two weeks after the scandal. As seen in Table 5, civil society organizations initially rejected this approved Regulation for not using the WHO/PAHO parameters, but then they accepted them and decided to advocate for the Manual’s design.

> “We reached a tacit agreement of accepting the Regulation with the parameters of Chile, right? Because we met at the College [of nutritionists] and we said: ‘If we fight the parameters of PAHO, this [the Regulation] will take even more time’; and, the fact that it takes longer, is going to imply that the other side – those against The Healthy Eating Law – would take advantage of it to create this kind of poorly made laws.” (Civil society organization’s representative, SOC-03)

#### 8.2.3. Proposed nutritional warnings in the period of designing the Regulation

None of the Regulation’s drafts specified a design for the nutritional warnings. According to our interviewees, during 2014 and 2015 advocates proposed the traffic lights design, which were the most well-known design at the time. However, based on the Ecuadorian experience, they discarded this design because it was not clear for consumers to decide which product was better and because it enabled the food industry to take advantage of their colors to merge them into colorful labels.

> “And we also had evidence that the other alternatives, like the traffic lights, were absolutely distracting and that they didn’t go to the essence of the problem. Let’s say, if we have a product with two orange balls, one green ball, and two red balls, who won? What is your conclusion? I mean, the interpretation is complex. And that was clear in Ecuador which was the first [country] to use this design.” (International organization’s representative, INT-01)

During 2015 onwards, the octagons design gained more support among advocates of the Law. Chile showed positive results in helping consumers decide (72) and not having severe effects in the food industry’s employment (73). Likewise, the Peruvian MoH conducted small studies using focus groups in different cities and found positive results with the octagons (14). Informants mentioned that the MoH tried to specify the octagons as the selected design for the nutritional warnings in the draft published in 2016. However, opponents at the PCM (e.g., MEF) argued that the Law did not mention using iconography for the warnings, so that the design should not be written. As shown in Table 5, to solve this disagreement, the Ministry of Justice was called to play as neutral party to decide, and it failed in favor of the opponents, which is why the published draft did not mention any specific design.

Throughout these years, opponents from the food industry and Congress were against the octagons and proposed the traffic lights or Guideline Daily Amounts (GDA) designs.

#### 8.2.4. Main challenges faced when trying to establish the nutritional warnings’ parameters in the Regulation

Main challenges during this period were: 1) the absence of parameters issued by WHO/PAHO until 2016, three years after the Law’s approval; 2) the strong opposition of the food industry, media, congressmen, and members of the PCM (e.g. Ministry of Economy, Ministry of Production) to the different parameters and nutritional warnings’ design proposed by the MoH; 3) the scarcity of local research to support the MoH’s decisions, mainly due to lack of funding and no alliances with researchers; 4) power asymmetries between those in favor and against (i.e., the food industry had plenty of resources to pay several lawyers to confront the Regulation’s drafts, whereas the MoH only had a small legal team involved in several other policies); and finally, 5) political will to assure the Regulation’s approval. In this regard, some interviewees mentioned that the former president Humala (2011–2016) supported the approval of the Law, but there was less consensus regarding his support to the Regulation:

> “When [Humala’s] government was finishing, President Humala questioned why if the Law begun in his period, the Regulation was not yet in place? He was also questioned about it. So, he said: ‘I want the Regulation now.’” (Decision maker at the MoH, DEC-01)
>
> “It’s perfectly known that the Law was approved, and President Humala turned around. He turned his back on it and did not want to move anything at all.” (International organization’s representative, INT-01)

### 8.3. In 2018, the Nutritional Warnings Manual is approved, defining the octagons as the selected design

#### 8.3.1. Summary of how the Manual was designed and approved

The MoH, in charge of designing the Manual, pre-published a first draft in August 2017, two months after the Regulation’s approval (48). The draft received several comments and queries, especially from the food industry, so the MoH assigned three persons to attend the comments aiming to approve it by November 2017. However, in September 2017, the Minister of Health was removed, and the next two ministers, ruling since September 2017 to April 2018, did not prioritized this policy.

> “You know how many times I have been told to put away the Manual? ‘Save your Manual, that’s never going to come out. Save it’. Then another [minister] came, another, another minister, and another director. And I took [the Manual] out again! (Laughs). People told me: ‘Don’t bother, it won’t have a chance, I don’t know why you’re insisting (…)’ And I looked for an audience to tell what the warning’s Manual was, yeah, until one [Minister] finally listened to me” (Decision maker at the MoH, DEC-04)

On the other hand, the new Congress, elected in July 2016 and with 60% of its congressmen from *Fuerza Popular*, the main opponent political party in the previous parliament, continued discussing bills to modify the Law and its Regulation. This was a very algid period for the policy, in which the MoH did not work on the Manual, and the Congress had enough votes to modify the Law and the warnings’ design. For instance, they proposed changing the octagons to traffic lights or GDA and replacing the nutritional warnings for nutritional tables at the back of the package. As shown in Table 3, six bills from different parties were prepared during 2017 (44-47, 49, 50) and then merged into one single document (Dictum). With majority of votes from *Fuerza Popular*, the Dictum was approved by the Congress on March 9^th^, 2018, and sent to the country President, Kuczynski, to either approve or veto the Dictum until April 9^th^, 2018.

On March 21^st^, however, a political crisis led President Kuczynski to his resignation (62), without having replied to the Dictum. On March 23^rd^, 2018, his vice-president, Vizcarra, assumed the presidency and days later he vetoed the Dictum, arguing that the MoH considered the octagons easier to understand and that the parameters proposed by the Congress (GDA) were based on a diet for an adult person (2000 kcal) and not children or adolescents (74).

> “[President] Vizcarra arrived and the first thing he heard was: ‘Sir, observe the Law’. And I’m sometimes pessimistic. I said: ‘How’s this man going to observe the Law, he’s just coming in, he doesn’t even know what it is, ahh, no, he is going to promulgate it, he won’t fight with the congressmen’. But [fortunately] he observed the Law!” (Civil society organization’s representative, SOC-01)

With President Vizcarra, the new minister of health resumed the Manual’s draft pre-published in 2017 and encouraged its approval. Meanwhile, the Congress continued preparing a new document to respond to the veto of President Vizcarra; however, and against all odds, they stopped their efforts, and the Manual was finally approved in June 2018 (14). Some informants believed that the Congress did not continue because *Fuerza Popular*, the main opponent political party was under public scrutiny and lost credibility due to denounces of corruption (75).

> “They [Fuerza Popular] cut off arms and legs to the Law, [but now] they wanted to implement their colored GDA. And they had the votes to do it. And [you can] remember that the president observed it [the Dictum]. That draft returned [to the Congress], and they [Fuerza Popular] had the votes to insist. If they would have insisted on [modifying] the Law, this would be history, we would not be talking about this right now.” (Congress member 2011-2016, POL-02)

Other interviewees argued that the confrontation between the MoH and the Congress reached the public opinion, and a dichotomy of health vs economy was created, where “the powerful food industry” was seen as only seeking to profit disregarding people’s health. Others added that a popular doctor who had a program in the biggest radio station (Dr. Elmer Huerta) was a key advocate to easily explain to the public the evidence supporting the octagons and to highlight the economic interests behind the proposals from the Congress. PAHO and civil society organizations were also strong supporters of the octagons.

> “Something that worked very well was having Elmer Huerta as a champion, right? (…) He [normally] tries to be fairly impartial on political issues; but, on this one, he wasn’t. And he was very clear in pointing out who were against the Law.” (Independent researcher, INV-03)

Once the Manual was approved, the nutritional warnings’ first implementation’s phase begun one year later, on June 17^th^, 2019; and its second phase, on September 17^th^, 2021.

#### 8.3.2. Proposed nutritional warnings in the period of designing the Manual

During these years, the nutritional warnings design was the main conflictive issue. Those in favor of the Law advocated for the octagons, arguing that they were easy to understand, and clearly enough to make rapid decisions (76). Opponents, on the contrary, proposed the traffic lights, GDA or a mix of both, arguing that consumers needed more information (i.e., quantity of nutrients) than solely short messages.

> “We ruled based on what the consumer law establishes, that a consumer must have all the appropriate information for decision making. So, that’s why we went for a mixed figure between the tables of nutritional values (…) and combine it with colors, or messages to inform if the minimum allowed was exceeded, according to the table established by the Ministry of Health.” (Congress member, 2016-2018, POL-03)

Based on qualitative studies conducted by the MoH, but mainly due to discussions at the PCM, the octagons’ design changed from the draft published in August 2017 to the Manual approved in June 2018. For instance, their colors changed from red to black, the label “Ministry of Health”, which was inside the red octagon, was removed from the warning, and the minimum package’s size that could be labelled changed from 20 cm^2^ to 50 cm^2^ (14).

> “The warnings’ Manual is basically about technical aspects, but there were also negotiations. No, no, they didn’t call it ‘negotiations’ but ‘decisions’ at the Senior Management [PCM], more political, let’s say, right? In other words, not everything was finally solved by the technical team. The technical team had a position, but the ultimate decision was at the Senior Management level” (Decision maker at the MoH, DEC-04)

#### 8.3.3. Main challenges faced when trying to establish the nutritional warnings’ design in the Manual

Main challenges in this period were 1) the strong power of the opponents at the Congress-level, risking the nutritional warnings policy; 2) the strong opposition to the octagons design at the PCM, leading to delays in the Manual’s approval and changes to the MoH’s proposed design; 3) the scarce political will of two health ministers to continue working on the Manual (Sept 2017 – Apr 2018); 4) political instability, which led to changes of the country President; 5) the limited locally-generated research to inform the MoH’s decisions regarding the warnings’ parameters and design, which was used by opponents as an argument to delegitimize the data from other countries that the MoH presented as evidence.

## 9. Why this policy was approved: Analysis using the Kaleidoscope Model

Following the Kaleidoscope Model (11), we conclude that the nutritional warnings *Reached the policy agenda* because the three main hypotheses were fulfilled. First, the nutritional warnings were part of a Law that targeted a relevant problem based on credible evidence and popular perception (Hypothesis 1). Unhealthy eating decisions among children and adolescents was unanimously seen as a relevant problem. Although opponents initially argued that the Law should focus on malnutrition and anemia rather than overweight and obesity, the Law’s aim of protecting children was strong enough to discourage more direct opposition. Likewise, even though during the Law’s design, opponents were against regulating publicity, which included the nutritional warnings’ aim of informing the population about processed food’s content, their narrative shifted later, when they no longer questioned this aim and instead focused on the “appropriate” warnings’ design.

Second, we identified well-defined focusing events (Hypothesis 2) that not only positioned the nutritional warnings in the policy agenda but helped it regain importance throughout the years. For example, the “expert consultation” organized by PAHO in 2011, where an initial set of parameters were proposed; the “Pura Vida” scandal in 2017, where supporters took the opportunity to reposition the Law and advocate for its Regulation; having the support of key health ministers who pursued the policy approval, despite oppositions at the Congress and PCM; having the Chilean experience of using octagons with promising results; and the political context during the Manual’s approval that reduced its opposition.

And third, there were strong individuals and institutions who supported the policy (Hypothesis 3) and were necessary along the years to maintain the policy’s position. Advocates came from diverse backgrounds, but mainly from the Congress, civil society organizations and PAHO. Importantly, having the support of most health ministers helped those at the MoH to continue working on the Regulation and Manual. Proponents from different backgrounds successfully joint together to push the policy, such as the MoH having close collaboration and technical guidance from PAHO; the alliance of many civil society organizations in a larger platform (“Healthy Eating Platform”); or maintaining key legal and technical personnel in the MoH working on the policy drafts, who gave continuity when ministers changed. Worth noting, even though supporters had some important discrepancies (See Table 5), they later agreed on joining efforts and move the policy forward.

Regarding the policy *Design*, in which the nutritional warnings’ design and parameters were defined, were possibly made mainly due to effective cost-benefit calculations (Hypothesis 6), in which advocates assessed their potential gains and losses with each design, as well as their own power and resources. In our case, the different proposed designs lead to several negotiations during the Regulation’s and Manual’s approval, yielding elements that weaken the regulations, but that ultimately allowed them to be approved. Importantly, the nutritional warnings’ purpose was to bring health gains at the population level (i.e., use of services’ savings, DALYs), especially for children and adolescents, which was used as a cost-effective argument to its approval.

On the contrary, the hypothesis of evidence and research as a key component to design a policy (Hypothesis 4) was the weakest one in this policy. One of the main limitations found throughout the process was the little evidence to back the proponents’ decisions about the parameters or warnings’ design, as well as the absence of alliances with researchers to generate local evidence or support the research efforts of advocates. Most studies were small-scale only and conducted by reduced groups at the MoH or civil society organizations (Table 3). Moreover, Peruvian entities did not allocate budget to support the conduction of robust studies to accompany this policy process.

Additionally, we found that the ideas and believes (Hypothesis 5) behind each key document along the process (e.g., among different Regulations’ drafts) substantially differed. For instance, the ideas of the first Regulation’s draft’s team were not acting against the food industry nor establishing parameters far different from the ones in the market; whereas the second Regulation’s draft team’s ideas were to legally back all their decisions due to the strong opposition faced from the food industry. Ideas and believes were also important in defining the warnings’ design, since advocates proposed the octagons, which have proven to be easy to read and understand (77), and better than other designs in helping consumers to choose the healthiest option among different products (78); in contrast to the traffic lights design, which misleadingly could favor a product high in a critic nutrient as healthy (77, 79).

Finally, for the *Adoption* stage, the three hypotheses of the model were found relevant for the policy approval. Government veto players (Hypothesis 8), such as Presidents and Health Ministers, were either in favor or neutral to the policy, but never against it, which enabled its continuation. However, powerful veto players at the Congress and within the PCM, related to the economic interests of the food industry, jeopardized the policy several times by trying to modify the Law or not approving the Regulation and Manual, especially during the political instability of 2016-2018. Even though neither the food industry nor civil society organizations had direct veto power, they were able to influence the policy process by press releases or media interviews, where they exposed their opinions and critics; or even by influencing decisions of congressmen and ministers. Worth noting, since this policy took several years, stakeholders changed along the years and some of them had greater interest and influence than others, as seen during the Manual’s period, where two Health Ministers did not make any efforts to move the policy forward. Others changed their stands, from being in favor to opposing, such as some congressmen during the approval of the Law.

Despite large disparities between opponents’ economic capacity in contrast to the scarce resources of the proponents, government stakeholders, such as Presidents and Health Ministries, had greater power than the food industry in terms of regulatory decisions (Hypothesis 7), since they were the ones in charge of designing the policy documents and ultimately, approving them. Importantly, this policy received public attention at different time points, which might have increased the power of proponents. For instance, the advocacy role played by a popular doctor in a radio station during the Manual’s approval helped raising the discussion to the public opinion, allowing proponents to gain more support. Lastly, even though supporters not always waited for propitious timings (Hypothesis 9) and generally had to wait for the long processes of negotiation or the existing calendars at the Congress and Ministers to position the policy, they did take advantage of some opportunities. The “Pura Vida” scandal is the most salient one, in which an international event received local attention and was used to advocate for the Regulation’s approval.

## IV. Discussion

This study describes, over the 2012-2018 period, the milestones and key stakeholders’ roles and stances during the Peruvian nutritional warnings policy design and approval, and analyzes the policy drivers that made it possible. The main milestones identified are the approvals of the three main policy documents: The Law 30021, its Regulation, and the Nutritional Warnings Manual. We found that the nutritional warnings were incorporated as part of the Law 30021, approved in 2013, after facing strong opposition from the media and food industry representatives and some congressmen. The nutritional warning parameters were differed to a further Regulation, which after four years and two policy drafts, was finally approved in 2017, soon after a mediatic scandal on a dairy product. This Regulation demanded a Manual to define the technical characteristics of the warnings, which after disagreements at the PCM and a parallel process of bills to modify the Law and its Regulation in the Congress, was approved in 2018. This Manual defined the black octagons as the warnings design and two implementation phases (June 2019 and September 2021) with different thresholds. Some relevant changes throughout this process were using more flexible parameters and taking out some specifications of the octagons, such as the label “Ministry of Health” or their use in small products. During this process, civil society organizations and PAHO were strong advocates for the policy.

Regarding key stakeholders’ roles and stances, we found that they came from different sectors and played relevant roles in favor or against the nutritional warnings policy along the years. The main opponent throughout the process was the food industry, which has proven to be a major opponent towards nutritional warnings policy in Latin American countries, such as Chile (80), Uruguay (7), Colombia (81, 82), and Brazil (83). The industry arguments in Peru were similar to those used in other countries when pursuing their nutritional warnings policies. For instance, critiques towards the evidence used by the promoters to support the parameters and warnings’ design, arguing that it was too weak (7), that the food industry’s proposals were not included (83), or that this policy did now showed reductions in obesity in other countries (82). Another common argument was the economic lost that the warnings’ implementation would bring to their countries, such as loss of jobs (7, 80, 83), rise of prices (7), or violations to international trade agreements (80, 83). None of these situations, however, occurred in the countries that have implemented the octagons (80). Importantly, beyond their arguments, we found that some powerful stakeholders were aligned to the food industry stance and acted against the warnings policy, such as congressmen and policymakers’ at ministries of economic sectors, as seen in Uruguay (7), Chile (80), and Colombia (82), which risked its approval and further implementation.

On the other hand, civil society organizations were vital to advocate for this policy by doing public appearances in TV and radio to inform about the policy and demanding its rapid approval and implementation. Similar to Colombia, these organizations promoted the bill to approve the nutritional warnings and the prohibition of foods with FoP labels to children and adolescents (82). Nonetheless, in Colombia, some health ministers were aligned with the food industry and against the policy (82); in clear contrast to Peru, where most health ministers were in favor of the policy, enabling its maintenance in the policy agenda and further approval. One important difference between Peru and other countries, such as Chile (84), Mexico (10), and Uruguay (7), have been the role of researchers and use of evidence to back the decisions of promoters. Whereas in Peru one of the main challenges across the years was the limited participation of researchers in the generation of local and tailored evidence, in other countries academics worked closely with policymakers and civil society organizations, supporting their arguments against opponents, and thereby easing the approval of their policies. Importantly, this reflection arises within a context and a policy environment where local support to science in the country is generally limited and, no less important, the spaces to promote the generation of local evidence to inform policies requires active promotion. These findings are important to anticipate in other settings where similar policies are yet to be pursued and where simultaneous support to the generation of local evidence will be essential to protect and progress with related whole of society prevention approaches.

Finally, another relevant finding is that beyond the technical contents needed to define the parameters and warnings’ design, the policy process of the nutritional warnings highly depended on political issues, such as the political will of key decision makers at the Congress and PCM and the political stability in the country. Indeed, the power of the opposition and their strong influence in the decisions made, continuously changed the policy contents and, potentially, their intended results. Thereby, this policy exemplifies the relevance of the political context during the design and approval of a health policy, which is usually ignored in academic studies (85).

## 10. Strengths and limitations

This study conveys the experiences of diverse and key supporters and opponents involved in the design and approval of the nutritional warnings’ policy in Peru, which allows to gain a deeper insight of how this process was and triangulate their different perspectives into a solid analysis. Likewise, having included the most relevant policy documentation and news about the nutritional warnings’ design and approval in our study enabled a better contextualization and analysis of our interviewees discourses. One limitation is that the elapsed time between the policy design and the interviewees, could potentially lead to a recall bias. However, their discourses were triangulated with other informants as well as with the policy documents and news.

## 11. Conclusion

This study presents the design process of the Nutritional Warnings policy in Peru, which was defined through three main documents: The Law 30021 (2013), its Regulation (2017), and the Nutritional Warnings Manual (2018). The stances of some stakeholders changed along the years, but in general, it was mainly supported by some congressmen, policymakers, civil society organizations, and PAHO; and usually opposed by the food industry and media representatives, as well as some civil society organizations and policymakers from the economic sector. The parameters and the warnings’ designs changed frequently across the years, with different proposals in each draft, mainly due to unclear evidence on which parameters and designs would be the best and difficulties to reach agreement among supporters and opponents. These changes led to important loses, such as using parameters more flexible than those proposed by PAHO or not using the octagons in small labels, however, they were also necessary negotiations to allow the approval of the policy.

Finally, based on the Kaleidoscope Model, we identified that the policy reached the political agenda because the problem it targeted (overweight and obesity in children and adolescents) proved to be relevant and secured the backing from powerful advocates within and outside the government, who took advantage of local and international focusing events to reposition the main policy documents at different stages. However, important weaknesses during the design were identified, especially the absence of official parameters and local studies to back decisions. Importantly, final versions of the documents were able to be shaped due to negotiations held by supporters, who considered the costs and benefits of their decisions and allowed editions that weakened the documents but enabled their completion. Even though the policy took several years before its implementation, it was ultimately adopted because government veto players (e.g., Health ministers, PCM) were mostly in favor of the policy, and despite having some relevant opposing players (e.g., Congressmen, PCM), the power of supporters proved to be stronger than that of their opponents. Acknowledging that the nutritional warnings policy has being implemented in different Latin American countries, these results will be useful for advocates to learn from the experience of Peru and anticipate potential difficulties and ways to overcome them.

## Data Availability

Data cannot be publicly shared because it contains potentially identifying information. Since we interviewed key stakeholders with specific and potentially unique knowledge about the nutritional warnings policy in Peru, the information they provided could ease their identification. Data requests could be sent to the corresponding author and they could be accessed on reasonable request

## Supplementary Material 1

### Code book

(K) = Codes aligned with the Kaleidoscope Model’s variables

#### 1. Law

1.1 (K) Champions during the Design

1.2 (K) Purpose of having nutritional warnings and/or designing a Law that includes the nutritional warnings

1.3 Documents (Law, bills)

1.3.1 Bills (2012)
1.3.2 Dictum (2013)
1.3.3 Approved Law (2013)
1.3.4 (K) Studies/reports conducted or used during the Law’s design
1.3.5 Experiences of warnings or legislation in other countries

1.4 Descriptions and opinions about the Law
1.4.1 About the parameters
1.4.2 About the nutritional warnings

1.5 Stakeholders’ postures/actions during the design and approval of the Law
  1.5.1 Food industry’s postures/actions
  1.5.2 Media (TV, newspapers, journalists)’s postures/actions
  1.5.3 Civil society organizations’ postures/actions
  1.5.4 Congress’s postures/actions
  1.5.5 Ministry of Health’s postures/actions
  1.5.6 Other government stakeholders’ postures/actions
  1.5.7 PAHO/WHO’s postures/actions
  1.5.8 Other stakeholders’ postures/actions

1.6 Difficulties for the design and/or approval of the Law

1.6.1 Specific difficulties: Lack of evidence
1.6.2 Specific difficulties: Power of the food industry
1.6.3 Specific difficulties: Power of opponents in the government
1.6.4 Specific difficulties: Power of other opponents

1.7 Enablers: Facts/actions that helped the Law to be designed and/or approved
  1.7.1 (K) Non-planned enablers. Facts that positioned the Law and nutritional warnings
  1.7.2 Intentional enablers. Actions/decisions taken so that the Law could be designed and/or approved

1.7.2.1 Intentional enablers. Actions/decisions that worked/were helpful

1.8 Recommendations or ideas that could enhance the Healthy Eating Law
  1.8.1 Explicit recommendations
  1.8.2 Proposals that got lost along the way
  1.8.3 What could have been done differently
  1.8.4 Critics

1.9 General assessments of the Law
1.10 Recommendations for other future policies (other than the Law and the octagons)
1.11 Law. Other topics

#### 2. Regulation of the Law

2.1 (K) Purpose of having a Regulation
2.2 Documents (Regulation, drafts)
  2.2.1 First Regulation’s draft (2014)
  2.2.2 Approval of parameters for sugar, sodium, and saturated fats (2015)
  2.2.3 Second Regulation’s draft (2016)
  2.2.4 Approval of parameters for trans fat (2016)
  2.2.5 Approved Regulation (2017)
  2.2.6 (K) Studies/reports conducted or used during the Regulation’s design

2.3 Descriptions and opinions about the Regulation and its drafts
  2.3.1 About the parameters of the First Regulation’s Draft (Local data)
  2.3.2 About the parameters of the Second Regulation’s Draft (PAHO)
  2.3.3 About the parameters of the approved Regulation (Chilean)

2.4 Stakeholders’ postures/actions during the design of the second Regulation’s draft
  2.4.1 Food industry’s postures/actions
  2.4.2 Civil society organizations’ postures/actions
  2.4.3 Ministry of Health’s postures/actions
  2.4.4 Other government stakeholders’ postures/actions
  2.4.5 PAHO/WHO’s postures/actions
  2.4.6 Other stakeholders’ postures/actions

2.5 Stakeholders’ postures/actions during the design of the approved Regulation
  2.5.1 Food industry’s postures/actions
  2.5.2 Civil society organizations’ postures/actions
  2.5.3 Ministry of Health’s postures/actions
  2.5.4 Other government stakeholders’ postures/actions
  2.5.5 PAHO/WHO’s postures/actions
  2.5.6 Other stakeholders’ postures/actions

2.6 Difficulties for the design and/or approval of the Regulation
  2.6.1 Specific difficulties: Lack of evidence
  2.6.2 Specific difficulties: Power of the food industry
  2.6.3 Specific difficulties: Power of opponents in the government
  2.6.4 Specific difficulties: Power of other opponents

2.7 Enablers: Facts/actions that helped the Regulation to be designed and/or approved
  2.7.1 (K) Non-planned enablers. Facts that positioned the Regulation
  2.7.2 Intentional enablers. Actions/decisions taken so that the Regulation could be designed and/or approved

2.7.2.1 Intentional enablers. Actions/decisions that worked/were helpful

2.8 Recommendations or ideas that could enhance the Regulation
  2.8.1 Explicit recommendations
  2.8.2 Proposals that got lost along the way
  2.8.3 What could have been done differently
  2.8.4 Critics

2.9 General assessments of the Regulation
  2.9.1 Reasons for the delay between the Law and the Regulation

2.10 Regulation: Other topics

#### 3. Nutritional Warnings’ Manual

3.1 (K) Purpose of having the Manual
  3.2 Documents (Manual, drafts)
    3.2.1 Manual’s draft (2017)
    3.2.2 Approved Manual (2018)
    3.2.3 (K) Studies/reports conducted or used during the Manual’s design
  3.3 Descriptions and opinions about the Manual and its draft
  3.4 Stakeholders’ postures/actions during the design of the Manual’s draft
    3.4.1 Ministry of Health’s postures/actions
    3.4.2 Other stakeholders’ postures/actions

3.5 Stakeholders’ postures/actions during the design of the approved Manual
  3.5.1 Food industry’s postures/actions
  3.5.2 Civil society organizations’ postures/actions
  3.5.3 Ministry of Health’s postures/actions
  3.5.4 Other government stakeholders’ postures/actions
  3.5.5 Other stakeholders’ postures/actions

3.6 Difficulties for the design and/or approval of the Manual
  3.6.1 Specific difficulties: Lack of evidence
  3.6.2 Specific difficulties: Power of the food industry
  3.6.3 Specific difficulties: Power of opponents in the government
  3.6.4 Specific difficulties: Power of other opponents

3.7 Enablers: Facts/actions that helped the Manual to be designed and/or approved
  3.7.1 (K) Non-planned enablers. Facts that positioned the Manual
  3.7.2 Intentional enablers. Actions/decisions taken so that the Manual could be designed and/or approved
    3.7.2.1 Intentional enablers. Actions/decisions that worked/were helpful

3.8 Recommendations or ideas that could enhance the Manual
  3.8.1 Explicit recommendations
  3.8.2 Proposals that got lost along the way
  3.8.3 What could have been done differently
  3.8.4 Critics

3.9 General assessments of the Manual
  3.9.1 Assessments of the octagons as a nutritional warning model
  3.9.1.1 Reasons why it was decided to use octagons
  3.9.2 Assessments of other warnings’ designs (Traffic light, GDA)

3.10 Manual: Other topics

4. Bills to amend the Law (and Regulation)
  4.1 Postures/actions of stakeholders in favor of the Law
  4.2 Postures/actions of opponents of the Law
  4.3 Power of opponents of the Law
  4.4 Enablers: Facts/actions that helped that the bills did not prosper

5. Comprehensive evaluations of all the documents

6. Comprehensive evaluations of all the stakeholders

## References

1. Popkin BM, Reardon T. Obesity and the food system transformation in Latin America. Obesity Reviews. 2018;19(8):1028–64.

2. Antiporta D, Miranda JJ. Ley de promoción de alimentación saludable: ¿jugando a la política con la salud de los niños? Revista Peruana de Medicina Experimental y Salud Publica. 2015;32:603-.

3. Organización Panamericana de la Salud. El etiquetado frontal como instrumento de política para prevenir enfermedades no transmisibles en la Región de las Américas. Washington, D.C: PAHO; 2020.

4. Instituto Nacional de Salud. Tendencia del sobrepeso y obesidad en niñas y niños de 5 a 9 años. INS; 2019.

5. Ley N° 30021. Ley de promoción de la alimentación saludable para niños, niñas y adolescentes, Ley N° 30021 (2013).

6. Villalobos Dintrans P, Rodriguez L, Clingham-David J, Pizarro T. Implementing a Food Labeling and Marketing Law in Chile. Health Systems & Reform. 2020;6(1):e1753159.

7. Ares G, Antúnez L, Cabrera M, Thow AM. Analysis of the policy process for the implementation of nutritional warning labels in Uruguay. Public Health Nutrition. 2021;24(17):5927–40.

8. Corvalán C. Evaluating new Chilean National Regulations on the Food Supply. Institute of Nutrition and Food Technology. Santiago: University of Chile, Santiago, Chile; Universidad Diego Portales, Santiago, Chile; University of North Carolina, USA; 2019.

9. James E, Lajous M, Reich MR. The Politics of Taxes for Health: An Analysis of the Passage of the Sugar-Sweetened Beverage Tax in Mexico. Health Systems & Reform. 2020;6(1):e1669122.

10. White M, Barquera S. Mexico Adopts Food Warning Labels, Why Now? Health Systems & Reform. 2020;6(1):e1752063.

11. Resnick D, Babu, S. C., Haggblade, S., Hendriks, S., & Mather, D.. Conceptualizing drivers of policy change in agriculture, nutrition, and food security: The kaleidoscope model. Washington DC: FPRI Discussion Paper. InternationalFood Policy Research Institute; 2015.

12. Haggblade S, Babu S. A user’s guide to the Kaleidoscope Model: Practical tools for understanding policy change 2017.

13. Decreto Supremo que aprueba el Reglamento de la Ley N° 30021, Ley de Promoción de la Alimentación Saludable, Decreto Supremo N° 017-2017-SA (2017).

14. Aprueban Manual de Advertencias Publicitarias en el marco de lo establecido en la Ley N° 30021, Ley de promoción de la alimentación saludable para niños, niñas y adolescentes, y su Reglamento aprobado por Decreto Supremo N° 017-2017-SA, Decreto Supremo N° 012-2018-SA (2018).

15. Bengtsson M. How to plan and perform a qualitative study using content analysis. NursingPlus Open. 2016;2:8–14.

16. Congreso de la República. Periodo Parlamentario 2011-2016. Sesión de instalación de la Junta Preparatoria 2011.

17. Organización Panamericana de la Salud. Recomendaciones de la Consulta de Expertos de la Organización Panamericana de la Salud sobre la promoción y publicidad de alimentos y bebidas no alcohólicas dirigida a los niños en la Región de las Américas.. Washington, DC: OPS; 2011.

18. Ollanta Humala Tasso asume presidencia de la República [press release]. 2011.

19. Foro Salud Perú. V Conferencia Nacional de Salud 17-11-11 (1ra parte). 2011.

20. Foro Salud. Declaración de la V Conferencia Nacional de Salud. Reforma del Estado en Salud, un imperativo ético y moral de inclusión social. V Conferencia Nacional de Salud; Peru 2011.

21. Proyecto de Ley 430/2011-CR, (2011).

22. Proyecto de Ley 775/2011-CR, (2011).

23. Proyecto de Ley 1038/2011-CR, (2011).

24. Midori De Habich asumió cargo como nueva Ministra de Salud [press release]. 2012.

25. Consejo Consultivo de Radio y Televisión CONCORTV. Análisis de la publicidad de alimentos no saludables en la televisión peruana. Lima: CONCORTV; 2012.

26. LEY 20606 sobre composición nutricional de los alimentos y su publicidad., (2012).

27. Aprueban el Reglamento que establece los parámetros técnicos sobre los alimentos y bebidas no alcohólicas procesados referente al alto contenido de azúcar, sodio y grasas saturadas y de la reducción gradual de grasas trans, Resolución Ministerial N° 321-2014/MINSA (2014).

28. Nombrar Ministro de Estado en el Despacho de Salud,al señor Aníbal Velásquez Valdivia, (2014).

29. Miranda Cipriano OR, Gómez Guizado GL, Munares García OF, Aquino Vivanco OS. Valores percentilares del contenido de azúcar, grasas y sodio en alimentos industrializados según etiquetado expendidos en lima. 2014.

30. Reglamento de etiquetado de alimentos procesados para consumo humano. Acuerdo Ministerial 5103. Registro Oficial Suplemento 318 (2014).

31. Organización Panamericana de la Salud. 53° Consejo Directivo. 66.a Sesión del Comité Regional de la OMS para las Américas. Washington, D.C.: OPS; 2014.

32. Proyecto de Ley que modifica diversos artículos de la Ley N° 30021, Ley de promoción de la alimentación saludable para niños, niñas y adolescentes, Proyecto de Ley 4343/2014-CR (2015).

33. Aprueban el Reglamento que establece los parámetros técnicos sobre los alimentos y bebidas no alcohólicas procesados referentes al contenido de azúcar, sodio y grasas saturadas, Decreto Supremo N° 007-2015-SA (2015).

34. Proyecto de Ley que modifica a la Ley 30021 “Ley de promoción de la alimentación saludable para niños, niñas y adolescentes”, Proyecto de Ley 4808/2015-CR (2015).

35. World Health Organization. WHO Regional Office for Europe nutrient profile model. World Health Organization: Geneva, Switzerland. 2015.

36. Ministerio de Salud. Reglamento de la Ley de Etiquetado de Alimentos Chile: MINSAL; 2015.

37. Colegio de Nutricionistas del Perú. Trome: Reclaman ley de comida sana. Lima CNP; 2016.

38. Decreto Supremo que aprueba el Reglamento de la Ley de promoción de alimentación saludable, Ley 30021, RM 524-2016/MINSA (2016).

39. Aprueban el Reglamento que establece el proceso de reducción gradual hasta la eliminación de las grasas trans en los alimentos y bebidas no alcohólicas procesados industrialmente, DS 033-2016-SA (2016).

40. Congreso de la República. Periodo Parlamentario 2016-2021. Sesión de instalación de la Junta Preparatoria. 2016.

41. Organización Panamericana de la Salud. Modelo de perfil de nutrientes de la Organización Panamericana de la Salud. Washington, DC OPS; 2016.

42. Pedro Pablo Kuczynski asume presidencia de la República [press release]. 2018.

43. Nombran Ministra de Salud, N° 164-2016-PCM (2016).

44. Proyecto de Ley 865/2016-CR, (2017).

45. Proyecto de Ley 1519/2016-CR, (2017).

46. Congreso de la República. Proyecto de Ley 1589/2016-CR. 2017.

47. Proyecto de Ley 1700/2016-CR, (2017).

48. Proyecto de Manual de Advertencias Publicitarias en el marco de lo establecido por la Ley N° 30021, RM 683/2017-MINSA (2017).

49. Proyecto de Ley 1959/2017-CR, (2017).

50. Proyecto de Ley 2036/2017-CR, (2017).

51. Proyecto de Ley del etiquetado de productos alimenticios (2017).

52. Fernando D’Alessio asumió conducción del Ministerio de Salud [press release]. 2017.

53. Español Ce. Así fue el camino de Pedro Pablo Kuczynski hasta la renuncia: los escándalos que lo sacudieron. CNN Latinoamérica 2018.

54. Conceden indulto y derecho de gracia por razones humanitarias a interno del Establecimiento Penitenciario Barbadillo, RS N° 281-2017-JUS (2017).

55. BBC News. ¿Es o no es leche?: la controversia por Pura Vida, el producto del gigante peruano de los lácteos Grupo Gloria cuya venta fue suspendida en Panamá. 2017.

56. ASPEC. NutriApp - Aplicativo con Toda La Información Nutricional de Productos. Canal N. 2017.

57. La República. Colegio de Nutricionistas realizará marcha contra reglamentación de Ley de Alimentación Saludable. 2017.

58. Accion Popular impuesta por el Colegio de Nutricionistas del Perú, Expediente 00344-2017-0-1801-SP-CI-02 (2017).

59. Ley que establece los parámetros técnicos sobre el contenido de azúcar, sodio y grasa saturada, y dispone el plazo para su observancia, Proyecto de Ley 2036/2017-CR (2018).

60. Oficio 061-2018-PR, (2018).

61. Nombran Ministro de Salud, RS N° 013-2018-PCM (2018).

62. Resolución Legislativa del Congreso por la que se acepta la renuncia del ciudadano Pedro Pablo Kuczynski Godard al cargo de Presidente de la República y se declara la vacancia de la Presidencia de la República, 008-2017-2018-CR (2018).

63. Andina. Martín Vizcarra juró como nuevo Presidente de la República. Andina Agencia peruana de noticias 2018.

64. Nombran Minsitra de Salud RS N° 091-2018-PCM (2018).

65. Redacción El Comercio. Martín Vizcarra: Fue un error mantener reserva a pedido de Keiko Fujimori. El Comercio. 2018.

66. Respuestas del Minsa a las consultas frecuentes sobre el Manual de Advertencias Publicitarias (2019).

67. Redacción El Comercio. Octógonos de advertencia: desde hoy rige su implementación en los productos. El Comercio 2019.

68. Dictamen. Comisión de Defensa del Consumidor y Organismos Reguladores de los Servicios Públicos, (2012).

69. Proyecto de Reglamento que establece los parámetros técnicos sobre los alimentos y bebidas no alcohólicas procesados referente al alto contenido de azúcar, sodio y grasas saturadas y de la reducción gradual de grasas trans, (2014).

70. Peñaflor-Guerra R, Sanagustín-Fons MV, Ramírez-Lozano J. Business Ethics Crisis and Social Sustainability. The Case of the Product “Pura Vida” in Peru. Sustainability. 2020;12(8):3348.

71. Proyecto de Reglamento Técnico que Regula Límites de Uso de Ácidos Grasos Trans, en Alimentos Elaborados Industrialmente, (2012).

72. Ministerio de Salud. Informe sobre evaluaciones de la Ley N° 20.606 sobre composición nutricional de los alimentos y su publicidad. Chile: Ministerio de Salud de Chile; 2021.

73. Paraje G, Colchero A, Wlasiuk JM, Sota AM, Popkin BM. The effects of the Chilean food policy package on aggregate employment and real wages. Food Policy. 2021;100:102016.

74. Oficio N° 061-2018-PR, (2018).

75. Fowks J. El Congreso peruano suspende al legislador Kenji Fujimori. El País. 2018.

76. Ministerio de Salud. Informe de evaluación de la implementación de la Ley sobre composición nutricional de los alimentos y su publicidad. Chile: MINSAL; 2018.

77. Valverde-Aguilar M, Espadín-Alemán CC, Torres-Ramos NE, Liria-Domínguez R. Preferencia de etiquetado nutricional frontal: octógono frente a semáforo GDA en mercados de Lima, Perú. Acta Médica Peruana. 2018;35:145–52.

78. Tórtora G, Machín L, Ares G. Influence of nutritional warnings and other label features on consumers’ choice: Results from an eye-tracking study. Food Research International. 2019;119:605–11.

79. Machín L, Aschemann-Witzel J, Curutchet MR, Giménez A, Ares G. Traffic Light System Can Increase Healthfulness Perception: Implications for Policy Making. Journal of Nutrition Education and Behavior. 2018;50(7):668–74.

80. Mialon M, Corvalan C, Cediel G, Scagliusi FB, Reyes M. Food industry political practices in Chile: “the economy has always been the main concern”. Globalization and Health. 2020;16(1).

81. Mialon M, Gaitan Charry DA, Cediel G, Crosbie E, Baeza Scagliusi F, Pérez Tamayo EM. “The architecture of the state was transformed in favour of the interests of companies”: corporate political activity of the food industry in Colombia. Globalization and Health. 2020;16(1).

82. Mialon M, Gaitan Charry DA, Cediel G, Crosbie E, Scagliusi FB, Perez Tamayo EM. ‘I had never seen so many lobbyists’: food industry political practices during the development of a new nutrition front-of-pack labelling system in Colombia. Public Health Nutrition. 2021;24(9):2737–45.

83. Mialon M, Khandpur N, Amaral Mais L, Bortoletto Martins AP. Arguments used by trade associations during the early development of a new front-of-pack nutrition labelling system in Brazil. Public Health Nutrition. 2021;24(4):766–74.

84. Reyes M, Garmendia ML, Olivares S, Aqueveque C, Zacarías I, Corvalán C. Development of the Chilean front-of-package food warning label. BMC Public Health. 2019;19(1).

85. Khan MM, Van den Heuvel W. The impact of political context upon the health policy process in Pakistan. Public Health. 2007;121(4):278–86.

